# Long-term immune persistence induced by two-dose BBIBP-CorV vaccine with different intervals, and immunogenicity and safety of a homologous booster dose in high-risk occupational population Secondary Study Based on a Randomized Clinical Trial

**DOI:** 10.1101/2022.06.22.22276690

**Authors:** Tian Yao, Xiaohong Zhang, Shengcai Mu, Yana Guo, Xiuyang Xu, Junfeng Huo, Zhiyun Wei, Ling Liu, Xiaoqing Li, Hong Li, Rongqin Xing, Yongliang Feng, Jing Chen, Lizhong Feng, Suping Wang

**Author notes:** Correspondence to: Suping Wang, School of Public Health, Center of Clinical Epidemiology and Evidence Based Medicine, Shanxi Medical University, Taiyuan, 030001, China, Lizhong Feng, Shanxi Provincial Center for Disease Control and Prevention, Shanxi Provincial Key Laboratory for major infectious disease response, Taiyuan, 030012, China, Jing Chen, Shanxi Provincial Center for Disease Control and Prevention, Shanxi Provincial Key Laboratory for major infectious disease response, Taiyuan, 030012, China, Yongliang Feng, School of Public Health, Center of Clinical Epidemiology and Evidence Based Medicine, Shanxi Medical University, Taiyuan, 030001, China. Contributed equally (Tian Yao, Xiaohong Zhang, Shengcai Mu). Joint supervisors (Yongliang Feng, Jing Chen, Lizhong Feng, Suping Wang).

## Abstract

**Background:** BBIBP-CorV vaccine with two doses and an interval of 3-4 weeks had been proved to have good immunogenicity and efficacy as well as an acceptable safety profile according to our initial research and other similar studies. Maintaining adequate neutralizing antibody levels is also necessary for long-term protection, especially in the midst of the COVID-19 pandemic. Our aim was to evaluate the immune persistence of neutralizing antibody elicited by BBIBP-CorV vaccines with day 0-14, 0-21 and 0-28 schedule, and assess the immunogenicity and safety of a homologous booster dose in the high-risk occupational population aged 18-59 years.

**Methods:** A total of 809 eligible participants, aged 18-59 years, were recruited and randomly allocated to receive BBIBP-CorV vaccine with day 0-14, 0-21 or 0-28 schedule respectively between January and May 2021 in Taiyuan City, Shanxi Province, China among the public security officers and the airport ground staff in initial study. In this secondary study, the responders (GMT ≥ 16) at day 28 after priming two-dose vaccine were followed up at months 3, 6 and 10 to evaluate the immune persistence of three two-dose schedules. At month 10, eligible participants of three two-dose schedules were received a homologous booster dose respectively (hereafter abbreviated as 0-14d-10m group, 0-21d-10m group and 0-28d-10m group), and followed up at day 28 post-booster to assess the safety and immunogenicity of the booster dose. The contents of follow-up included the blood samples, oropharyngeal/nasal swabs, and adverse reactions collection. The main outcomes of the study included geometric mean titers (GMT) of neutralizing antibody to live SARS-CoV-2, the positive rates of different criteria and the constituent ratio of GMT of neutralizing antibodies at different follow-up point. Meanwhile, we explored the kinetics of antibody levels of different vaccination regimens by generalized estimating equations (GEE) and used exponent curve model to predict the duration of maintaining protected antibody after the booster dose. We also determined predictors of maintaining protected antibody level within 10 months after the second dose by Cox proportional hazards regression model and nomogram. The trial was registered with ChiCTR.org.cn (ChiCTR2100041705, ChiCTR2100041706).

**Results:** The number of 241, 247 and 256 responders (GMT ≥ 16) at day 28 after two-dose BBIBP-CorV vaccine in 0-14d, 0-21d and 0-28d schedule were followed-up at months 3, 6, and 10 for immune persistence evaluation. At month 10, a total of 390 participants were eligible and received a booster dose with 130 participants in the 0-14d-10m, 0-21d-10m and 0-28d-10m group respectively, of whom 74.1% (289/390) were male, with a mean age of 37.1±10.3 years. The GMT of neutralizing antibody in 0-28d-10m and 0-21d-10m group were significantly higher than 0-14d-10m group at month 3 (GMT: 71.6 & 64.2 vs 46.4, *P*<0.0001), month 6 (GMT: 47.1 & 42.8 vs 30.5, *P* < 0.0001) and month 10 (GMT: 32.4 vs 20.3, *P* < 0.0001; 28.8 vs 20.3, *P*=0.0004) after the second dose. A sharply decrease by 4.85-fold (GMT: 94.4-20.3), 4.67-fold (GMT: 134.4-28.8) and 4.49-fold (GMT: 145.5-32.4) was observed from day 28 to month 10 after the second dose in 0-14d-10m, 0-21d-10m and 0-28d-10m group, respectively, and they had similar decline kinetics (*P*=0.67). At 28 days after booster dose, a remarkable rebound in neutralizing antibody (GMT: 246.2, 277.5 and 288.6) were observed in three groups, respectively. Notably, the GMT after booster dose was not affected by priming two-dose schedule. The predictive duration of neutralizing antibody declining to the cutoff level of positive antibody response may be 18.08 months, 18.83 months and 19.08 months after booster dose in three groups, respectively. Long-term immune persistence within 10 months after the second dose was associated with age<40, female, and history of influenza vaccination. All adverse reactions were mild after the booster injection. None of the participants were infected SARS-CoV-2 during the trial period.

**Conclusions:** The priming two-dose BBIBP-CorV vaccine with 0-28 days and 0-21 days schedule could lead a longer persistence of neutralizing antibody than 0-14 days schedule. Maintaining long-term immune persistence was also associated with age<40, female, and history of influenza vaccination. Regardless of priming two-doses vaccination regimens, a homologous booster dose led to a strong rebound in neutralizing antibody and might elicit satisfactory persistent immunity.

## Background

Spread of severe acute respiratory syndrome coronavirus 2 (SARS-CoV-2) infections has led to a substantial threat to public health worldwide. Globally, as of 20, June 2022, there have been 536.59 million confirmed cases of COVID-19, including 6.32 million deaths, according to the World Health Organization (WHO) COVID-19 Dashboard [1]. Vaccination is the most effective approach to the long-term strategy of COVID-19 prevention and control. WHO listed the BBIBP-CorV vaccine for emergency use on 7 May 2021, giving the approval for this vaccine to be rolled out globally [2]. Our initial research and other similar studies indicated that a longer interval (21 days and 28 days) between the first and second BBIBP-CorV vaccination produced higher neutralizing antibody levels compared with a shorter interval schedule (0-14 day) [3, 4].

Adequate neutralizing antibody levels could serve as a correlation of protection for vaccines against SARS-CoV-2 in humans [5-7], but immune protection from infection may wane with time as neutralizing antibody levels decline [6]. Whereas vaccines induce durable T and B cell memory to SARS-CoV-2 [8, 9], strong evidence revealed the crucial protective role for neutralizing serum antibodies[6] due to their ability to block the viruses from entering the host cells directly [10]. So, maintaining adequate neutralizing antibody levels is more necessary for long-term protection, especially during the midst of the COVID-19 pandemic.

Recent studies indicated that binding or neutralizing SARS-CoV-2 antibodies elicited by multiple types of vaccines (mRNA vaccines, adenovirus-vectored vaccines, and inactivated vaccines et al) declined to varying degrees over time after priming full-schedule vaccination, and showed substantial descent by 6 to 12 months [11-17]. Meanwhile, age, sex, comorbidities and immunosuppressed condition had an impact on neutralizing antibody loss [17-19]. However, lasting immune response can certainly be affected by spacing between the doses [20], and a complete picture of the kinetics of long-term immune persistence elicited by priming two-dose BBIBP-CorV vaccine with different vaccination interval is not yet available. Besides, the fading immune response and the emergence of the Omicron (B.1.1.529) variant had raised concerns about the booster dose. Recent research revealed that a booster dose of different type of SARS-CoV-2 vaccine could induce remarkable high neutralizing antibody levels [21-23]. Nevertheless, the safety and immunogenicity of a booster dose with different priming two-dose BBIBP-CorV vaccination regimens are yet to be thoroughly evaluated, especially in the high-risk occupational population.

Consequently, this paper assessed immune persistence of priming two-dose BBIBP-CorV inactivated vaccines, and evaluated the immunogenicity and safety of a homologous booster dose among three different vaccination regimens based on our initial study. Meanwhile, we explored the kinetics of neutralizing antibody levels of different vaccination regimens and determined predictors of maintaining protected antibody levels, and constructed an exponent curve model to predict neutralizing antibody decay after booster dose.

## Methods

### Study design and participants

Between on January, 2021 and May, 2021, a randomized, controlled phase IV clinical trial of BBIBP-CorV vaccine with three two-dose schedules was conducted in Taiyuan City, Shanxi Province, China. Public security officers and airport ground staff aged 18-59 years, without previous SARS-CoV-2 vaccination and infection were eligible for enrollment. A complete list of inclusion and exclusion criteria can be found in a previous publication [3] (see protocol 2 p 1). After initial study, participants who were neutralizing antibody positive [geometric mean titer (GMT) of 16, positive response was defined as at least a 4-fold increase of neutralizing antibody titers over baseline (4)] at day 28 after priming two-dose vaccine were followed-up extended to November, 2021 (10 month after priming two-dose vaccination) for assessing the immune persistence of three two-dose schedules of BBIBP-CorV vaccine. Subsequently, on Nov 26, 2021, some participants who met the inclusion and exclusion criteria were eligible to receive the booster injection for evaluating the safety and immunogenicity of booster dose. Inclusion criteria included that (1) No an unacceptable adverse event; (2) No SARS-CoV-2 infection during the follow-up period; (3) Completed the initial two doses of vaccination; (4) be able and willing to receive the booster dose injection. Exclusion criteria included that (1) A COVID-19 vaccine other than the experimental vaccine was used during the study period; (2) Acute or new chronic disease occurs during follow-up; (3) other vaccination history within 14 days before vaccination; (4) being pregnant or breastfeeding before vaccination. The detailed inclusion and exclusion criteria are available at protocol 2 p 5.

Written informed consent was obtained from all participants before the enrolment in the initial study. The clinical trial protocol for the study was approved by the Ethics Committee of Shanxi Provincial Center for Disease Control. The study was done in accordance with the Declaration of Helsinki and Good Clinical Practice. The trial was registered with ChiCTR.org.cn (ChiCTR2100041705, ChiCTR2100041706).

### Procedures

In initial study, eligible participants were recruited and randomly allocated (1:1:1) to receive BBIBP-CorV vaccine of 4µg with three two-dose schedules on days 0 and 14, 0 and 21, or 0 and 28, respectively.

The responders at day 28 after priming two-dose vaccine were followed up at months 3, 6 and 10 in this secondary study to evaluate the immune persistence of the second dose. At month 10 after the second dose, eligible participants of three two-dose schedules were received the homologous booster dose respectively (hereafter abbreviated as 0-14d-10m group, 0-21d-10m group and 0-28d-10m group), and followed up at day 28 post-booster to assess the safety and immunogenicity of the booster dose. The vaccines were administered intramuscularly in the deltoid region of the upper arm with a dosage of 4µg. The vaccines used in this study were inactivated vaccine (Vero Cell) developed by the Beijing Institute of Biological Products (Beijing, China). The contents of follow-up included the blood samples, oropharyngeal/nasal swabs, and adverse reactions collection. Fig. 1 shows the study protocol.

**Figure 1:**
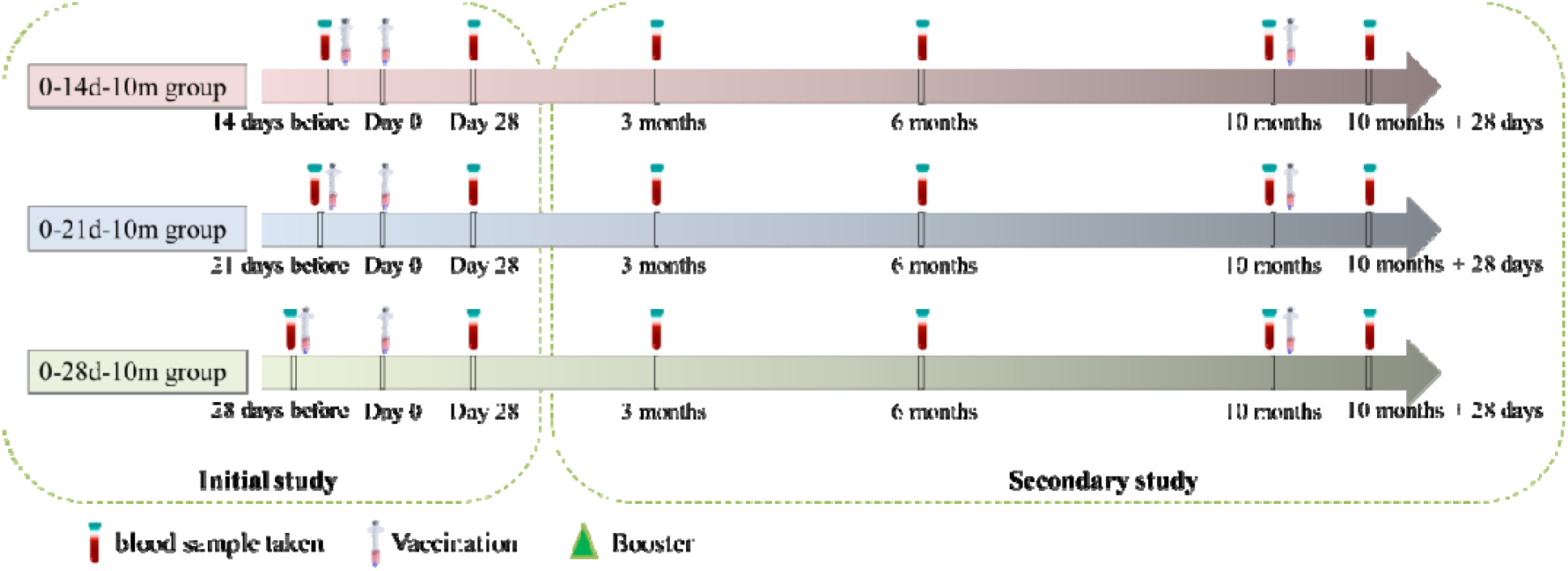
Trial process timeline.

### Safety assessment

Safety information after the booster dose was obtained by the same methods as for priming two doses [3]. Participants were observed for 30 minutes in the observation room after booster dose vaccination for any acute reactions. Participants were instructed to record local and systemic reactions daily for 7 days on diary cards. Solicited local adverse events included pain at the injection site, induration and swelling, and systemic adverse events included diarrhea and dysphagia. For days 8-28, participants spontaneously reported any unsolicited adverse events including local reactions (rash) and Systemic reactions (cough and headache). Adverse reactions were graded according to the Guidelines for Adverse Event Classification Standards for Clinical Trials of Preventive Vaccines by the National Medical Products Administration (version 2019) [24].

### Laboratory methods

Oropharyngeal/nasal swabs for RT-PCR (reverse transcriptase-polymerase chain reaction) testing were collected from all participants at each follow-up point. The neutralizing antibody to live SARS-CoV-2 (strain 19nCoV-CDC-Tan-Strain 05 [QD01]) were quantified using a micro cytopathogenic effect assay at 3, 6 and 10 months after the second dose, and at day 28 after the booster dose. Blood samples taken at baseline and at day 28 after the second dose had been tested previously [3]. The lower limit of detection was 4 for the neutralizing antibody test. We defined positive antibody response as a titer of 16 or greater for neutralizing antibody levels to infectious SARS-CoV-2. We assessed the positive rates of neutralizing antibodies with different criteria (GMT ≥ 16, 32, 64, 128 or 256).

### Outcomes

The primary immunological endpoints of the initial trials can be found in an initial publication [3]. Here, we report the results of secondary and exploratory immunological endpoints. Secondary immunogenic endpoints included GMT of neutralizing antibody to live SARS-CoV-2, the positive rates of different criteria and the constituent ratio of GMT of neutralizing antibodies (GMT 16-31,GMT 32-63, GMT 64-127,GMT 128-255 and GMT ≥ 256) on 3, 6 and 10 months after two dose and at day 28 after booster dose. Exploratory immunogenic endpoints included the kinetics of antibody levels of different vaccination regimens, predictors of maintaining protected antibody levels within 10 months after two doses of vaccination and the predictive duration of booster-induced neutralizing antibody declining to the cutoff level of positive antibody response. The primary safety endpoints included adverse events within 7 days after booster dose. Secondary safety endpoints were any adverse events within 28 days after the booster dose vaccinations across the three groups.

### Statistical analyses

We followed up participants who were antibody positive at day 28 after priming two-dose vaccine. During the follow-up, if the neutralizing antibody response of participants was negative, they will no longer be followed up, and the antibody response at each time point thereafter was negative by default. When the previous observed titer was positive and still positive in the next follow-up, we imputed the intermediate missing value as a positive response and the antibody titers were filled according to the next follow-up. If neutralizing antibody was positive at the previous follow-up and negative at the next follow-up, the intermediate missing value would not be filled. The imputed nonresponses and responses and the observed neutralizing antibody levels were together referred to as complete case data (see appendix p 2-3).

The complete case data was used to calculate the GMT, positive rates and the constituent ratio of neutralizing antibodies at each follow-up point. Safety endpoints were presented descriptively as frequencies (%) per group. Analysis of Variance (ANOVA) was used to analyze neutralizing antibody levels of different groups which were log-transformed. Chi-square test or Fisher precision test was used to Categorical data. The kinetics of neutralizing antibody levels, involving group, time, and interaction between group and time, were analyzed by generalized estimating equations (GEE), and the paired *t*-test was used to analyze the differences in neutralizing antibody levels among different follow-up point [*P* value for statistical significance was 0.008 (0.05/6)]. Cox proportional hazards regression model was used to assess the influencing factors associated with maintaining of neutralizing antibody levels during 10 months after the second dose. The nomogram was constructed to predict the long-term immune persistence mainly based on the results of the Cox proportional hazards regression model. The discrimination ability was assessed using the Concordance indexes (C-index). C-index vary from 0.5 to 1.0 and C-index values greater than 0.7 suggest a reasonable estimation.

An exponent curve model was used to predict the neutralizing antibody levels after the booster dose, and the general form of exponential curve equation is Ŷ = *k* + *α* exp (*b*X). The position, direction and curvature of a curve are determined by the sign and magnitude of constant terms and parameters *α* and *b*, respectively. The actual antibody levels (Y) over time (X) of follow-up from 28 days (calculated according to 1 month) to 10 months after two doses were used to construct the model. After the actual values were transformed into logarithm, reciprocal or square root, the curve equation was linearized, and the least square method was used to fit the optimal exponential model of vaccine immune persistence (see appendix p 13-15). Based on the model, the antibody attenuation after enhancement was predicted according to the actually observed neutralizing antibody titer decay after two doses. Other values of *P*<0.05 were considered statistically significant. The nomogram was performed using R software, version 4.1.3. Other statistical computations were performed using SAS version 9.4 and graphing performed using GraphPad Prism 9.0.0.

## Result

### Participants’ characteristics at baseline

A total of 809 participants were enrolled and randomly assigned to the 0-14d (n=270), 0-21d (n=270) and 0-28d (n=269) schedule, respectively. The responders (256, 247 and 241 participants) at day 28 after priming two-dose vaccination were followed-up at months 3, 6, and 10 for immune persistence evaluation, respectively. At month 10 after second dose, a total of 390 participants were eligible and received a booster dose, with 130 participants in the 0-14d-10m, 0-21d-10m and 0-28d-10m group respectively, of whom 74.1% (289/390) were male and 25.9% (101/390) were female, with a mean age of 37.1±10.3 years. There were 122, 120 and 112 participants in three groups completed blood sampling at 28 days after the booster dose (Fig. 2; Table 1). The demographic characteristics were broadly similar between the participants who received booster dose of vaccine and no vaccination. There was no difference in demographics characteristics among the three groups at each follow-up point (see appendix p 4-8).

**Table 1:**
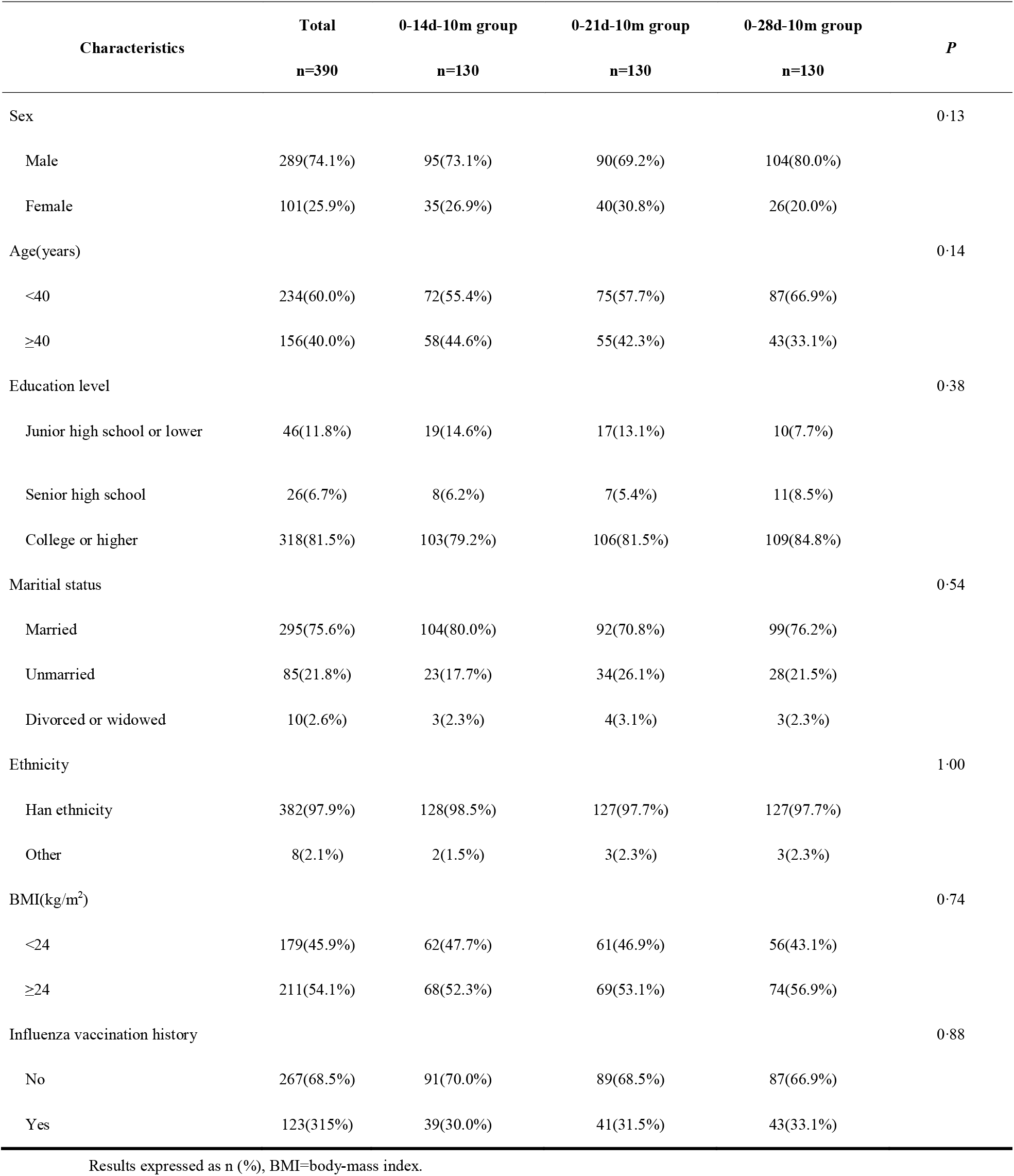
Demographic and Behavioral Characteristics of High-risk Occupational Population who received the Booster Dose.

| Characteristics | Total<br>n=390 | 0-14d-10m group<br>n=130 | 0-21d-10m group<br>n=130 | 0-28d-10m group<br>n=130 | *P* |
| --- | --- | --- | --- | --- | --- |
| Sex |  |  |  |  | 0.13 |
| Male | 289(74.1%) | 95(73.1%) | 90(69.2%) | 104(80.0%) |  |
| Female | 101(25.9%) | 35(26.9%) | 40(30.8%) | 26(20.0%) |  |
| Age(years) |  |  |  |  | 0.14 |
| <40 | 234(60.0%) | 72(55.4%) | 75(57.7%) | 87(66.9%) |  |
| ≥40 | 156(40.0%) | 58(44.6%) | 55(42.3%) | 43(33.1%) |  |
| Education level |  |  |  |  | 0.38 |
| Junior high school or lower | 46(11.8%) | 19(14.6%) | 17(13.1%) | 10(7.7%) |  |
| Senior high school | 26(6.7%) | 8(6.2%) | 7(5.4%) | 11(8.5%) |  |
| College or higher | 318(81.5%) | 103(79.2%) | 106(81.5%) | 109(84.8%) |  |
| Marital status |  |  |  |  | 0.54 |
| Married | 295(75.6%) | 104(80.0%) | 92(70.8%) | 99(76.2%) |  |
| Unmarried | 85(21.8%) | 23(17.7%) | 34(26.1%) | 28(21.5%) |  |
| Divorced or widowed | 10(2.6%) | 3(2.3%) | 4(3.1%) | 3(2.3%) |  |
| Ethnicity |  |  |  |  | 1.00 |
| Han ethnicity | 382(97.9%) | 128(98.5%) | 127(97.7%) | 127(97.7%) |  |
| Other | 8(2.1%) | 2(1.5%) | 3(2.3%) | 3(2.3%) |  |
| BMI(kg/m<sup>2</sup>) |  |  |  |  | 0.74 |
| <24 | 179(45.9%) | 62(47.7%) | 61(46.9%) | 56(43.1%) |  |
| ≥24 | 211(54.1%) | 68(52.3%) | 69(53.1%) | 74(56.9%) |  |
| Influenza vaccination history |  |  |  |  | 0.88 |
| No | 267(68.5%) | 91(70.0%) | 89(68.5%) | 87(66.9%) |  |
| Yes | 123(31.5%) | 39(30.0%) | 41(31.5%) | 43(33.1%) |  |

Results expressed as n (%), BMI=body-mass index.

**Figure 2:**
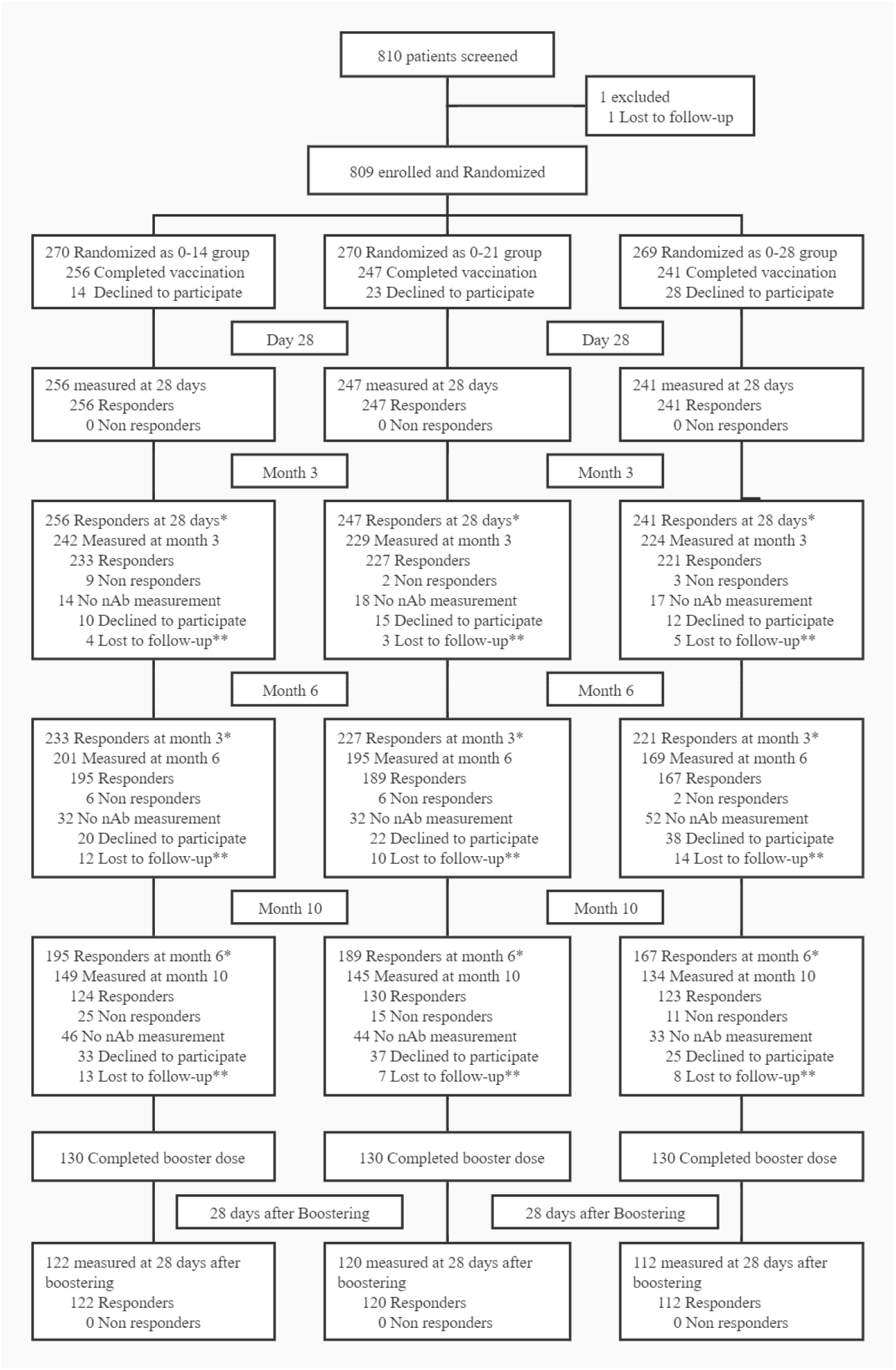
Flow of participants in a study of the inactivated SARS-CoV-2 vaccine in high-risk occupational population. * Participants who were antibody positive (GMT≥16) could be followed up **Lost to follow-up including transfer, business trip or physical discomfort nAb: neutralizing antibody

### GMT , positive rates and constituent ratio of SARS-CoV-2 neutralizing antibodies at each follow-up point in three groups

The GMT of neutralizing antibodies at month 3 after the second dose was 46.4 [95% confidence interval (*CI)*: 41.4∼52.0] in the 0-14d-10m group, which was significantly lower than 64.2 (95%*CI:* 58.8∼70.1) in the 0-21d-10m group (*P*<0.0001), and 71.6 (95%*CI:* 65.7∼78.0) in the 0-28d-10m group (*P*<0.0001). The GMT of neutralizing antibodies at month 6 after the second dose reduced to 30.5 (95%*CI:* 27.0∼34.5) in the 0-14d-10m group, 42.8 (95%*CI:* 39.2∼46.7) in the 0-21d-10m group (*P*<0.0001 vs 0-14d-10m group), and 47.1 (95%*CI:* 42.2∼52.6) in the 0-28d-10m group (*P*<0.0001 vs 0-14d-10m group). At month 10 after the second dose, the GMT of neutralizing antibodies declined to 20.3 (95%*CI:* 17.4∼23.6) in the 0-14d-10m group, 28.8 (95%*CI:* 25.5∼32.6) in 0-21d-10m group (*P=*0.0004, vs 0-14d-10m group), and 32.4 (95%*CI:* 28.7∼36.6) in the 0-28d-10m group (*P*<0.0001, vs 0-14d-10m group). (Fig. 3; see appendix p 9). Results of post-booster immunogenicity analysis showed that the GMT on day 28 after booster vaccination increased to 246.2 (95%*CI:* 222.6-269.7), 277.5 (95%*CI:* 248.9-306.0) and 288.6 (95%*CI:* 256.9-320.3) in 0-14d-10m, 0-21d-10m and 0-28d-10m group, respectively (Fig. 3). There was no significant difference in the GMT of post-booster neutralizing antibodies among the three groups (*P*>0.05) (see appendix p 9).

**Figure 3:**
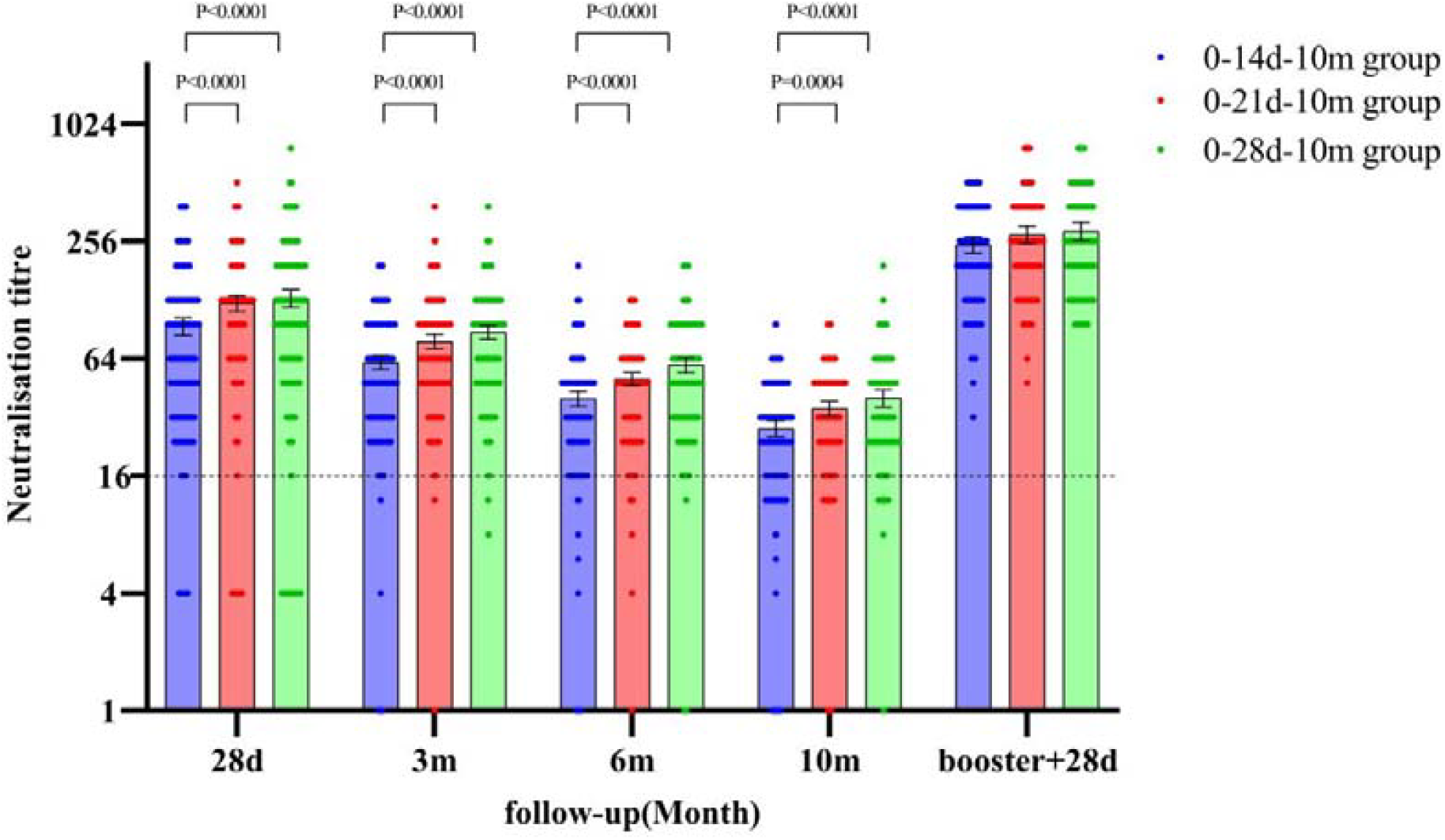
Neutralizing antibody titers of SARS-CoV-2 vaccine at each follow-up point in high-risk occupational population. Dots are reciprocal neutralizing antibody titers for individuals in the different groups. The bars represent GMT, and the error bars indicate the 95% *CI*. The dotted horizontal line represents the positive response threshold. GMT=geometric mean titer.

From month 3 to 10 after the second dose, the positive rates of neutralizing antibodies decreased from 96.3% (237/246) to 76.7% (132/172) in the 0-14d-10m group, from 99.2% (233/235) to 86.5% (147/170) in the 0-21d-10m group and from 98.7% (228/231) to 89.8% (141/157) in the 0-28d-10m group, respectively. At 28 days after booster dose, positive rates in all vaccination groups markedly increased in 100.0%. (Fig. 5; see appendix p 9). Details of the positive rates of neutralizing antibodies with different criteria at each follow-up point can be found in the appendix p 9.

**Figure 4:**
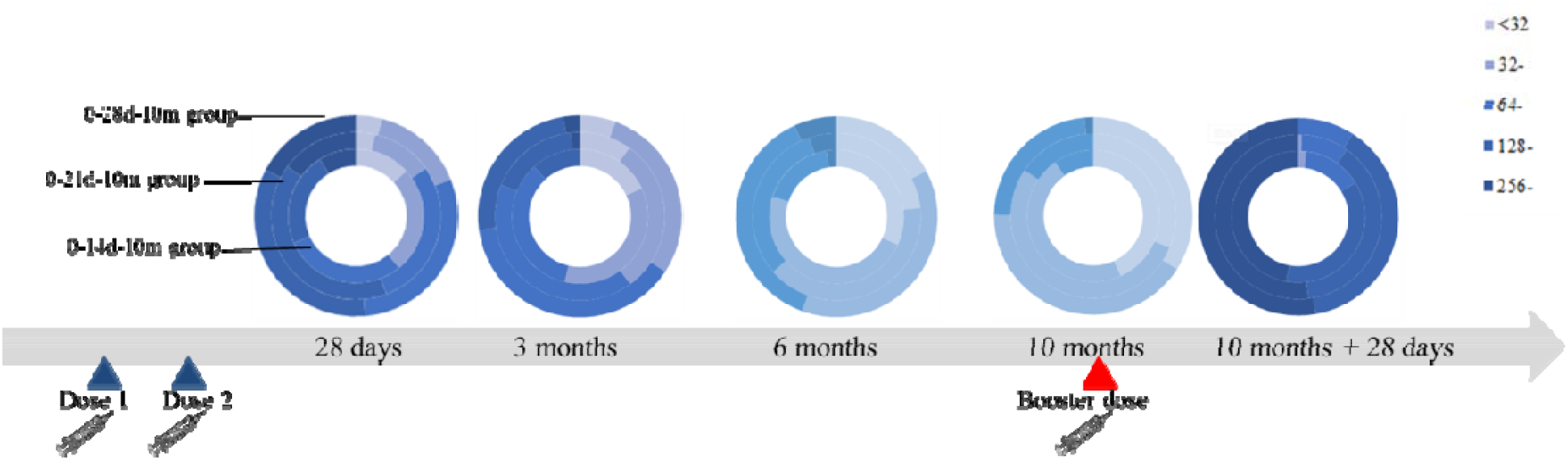
Constituent ratio of neutralizing antibodies at each follow-up point in high-risk occupational population.

**Figure 5:**
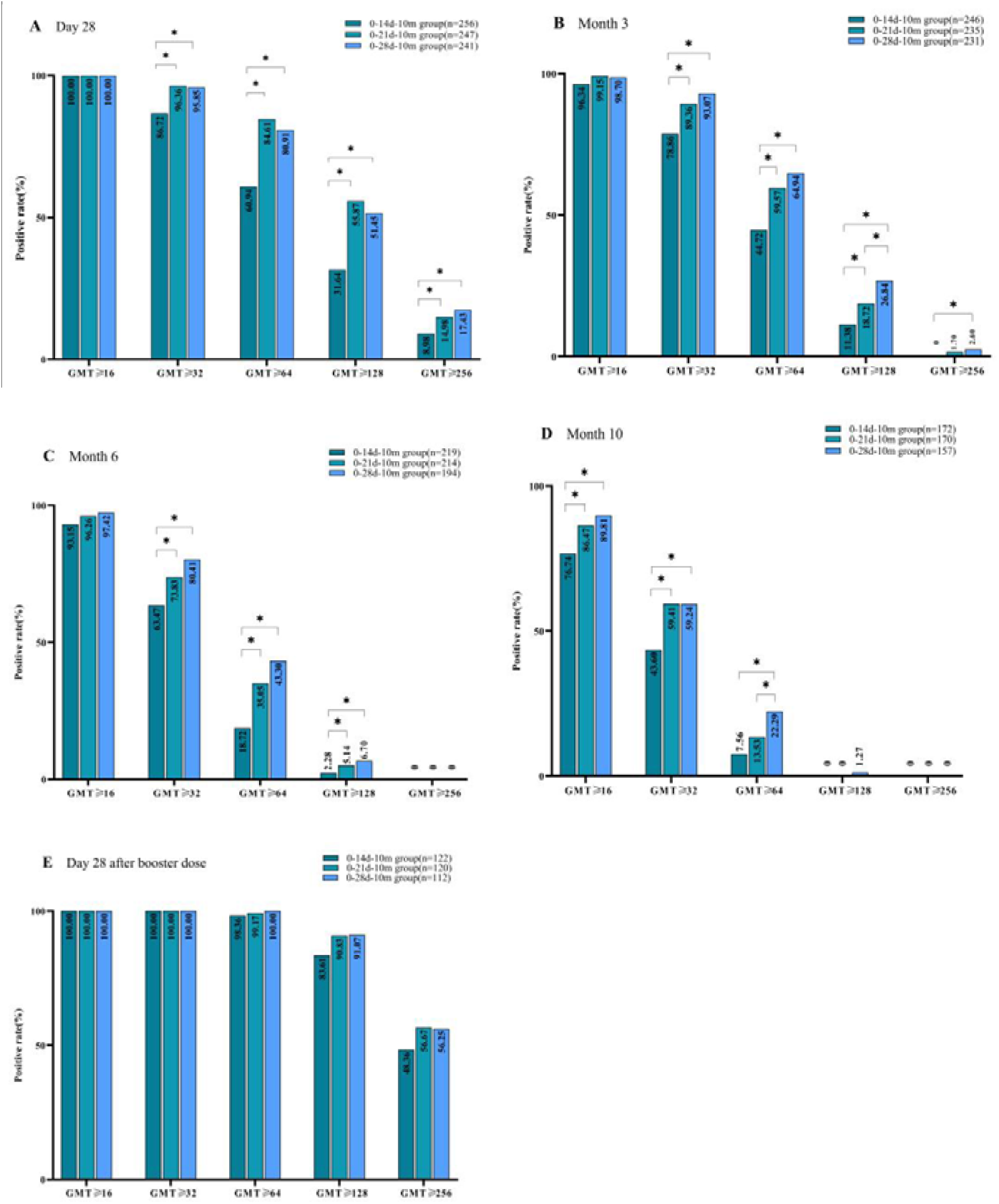
Positive rate within different criteria at each follow-up point in high-risk occupational population.

On day 28 after the second dose, the constituent ratio of neutralizing antibodies was higher at the GMT 64-127 (29.3%, 75/256) in the 0-14d-10m group, while the constituent ratio of neutralizing antibodies in the 0-21d-10m and 0-28d-10m group was higher at the GMT of 64-127 (28.8%, 71/247 ; 29.5%, 71/241) and 128-255 (40.9%, 101/247; 34.0%, 82/241), respectively. However, on 10 months after the second dose, the constituent ratio of neutralizing antibodies was sharply decrease at the GMT 64-127 (9.9%, 13/132) in the 0-14d-10m group, while the constituent ratio of neutralizing antibodies in the 0-21d-10m and 0-28d-10m groups was remarkably decrease at the GMT of 64-127 (15.7%, 23/147; 23.4%, 33/141) and 128-255 (0.0%, 0/147; 1.4%, 2/141), respectively. On day 28 after booster vaccination, a strangely rebound in the constituent ratio of neutralizing antibodies was observed at the GMT 128-255 (35.2%, 43/122; 34.2%, 41/120; 34.8%, 39/112) and GMT≥256 (48.4%, 59/122; 56.7%, 68/120; 56.3%, 63/112) in three groups, respectively (Fig. 4, see appendix p 10).

### Kinetics of SARS-CoV-2 neutralizing antibodies in three groups and predictive antibody decay after booster dose

From day 28 to month 10 after second dose, a substantial reduction of SARS-CoV-2 neutralizing antibodies by 4.85-fold (GMT: from 98.4 to 20.3, *P*<0.0001), 4.67-fold (GMT: from 134.4 to 28.8, *P*<0.0001) and 4.49-fold (GMT: from 145.5 to 32.4, *P*<0.0001) were observed in 0-14d-10m group, 0-21d-10m group and 0-28d-10m group, respectively. In the whole process, the GMT of neutralizing antibodies decreased more rapidly of 2.12-fold (GMT: from 98.4 to 46.4, *P*<0.0001), 2.09-fold (GMT: from 134.4 to 64.2, *P*<0.0001) and 2.03-fold (GMT: from 145.5 to 71.6, *P*<0.0001) from days 28 to months 3, but slowed thereafter in three groups, respectively (Table 3). The kinetic profile was similar among three groups by GEE (*P*=0.67) (Fig. 6; Table 2). On day 28 post-booster-vaccination, the GMT of neutralizing antibodies increased from pre-boosting levels (10 months after the second dose) by 12.16-fold (GMT: from 20.3 to 246.2, *P*<0.0001) in 0-14d-10m group, by 9.64-fold (GMT: from 28.8 to 277.5, *P*<0.0001) in the 0-21d-10m group and by 8.91-fold (GMT: from 32.4 to 288.6, *P*<0.0001) in the 0-28d-10m group, respectively. Correspondingly, booster vaccination led to 2.50-fold (GMT: from 98.4 to 246.2, *P*<0.0001), 2.06-fold (GMT: from 134.4 to 277.5, *P*<0.0001) and 1.98-fold (GMT: from 145.5 to 288.6, *P*<0.0001) increases from day 28 after the second dose to day 28 post-booster in the GMT of neutralizing antibodies in three groups, respectively (Table 3).

**Table 2:**
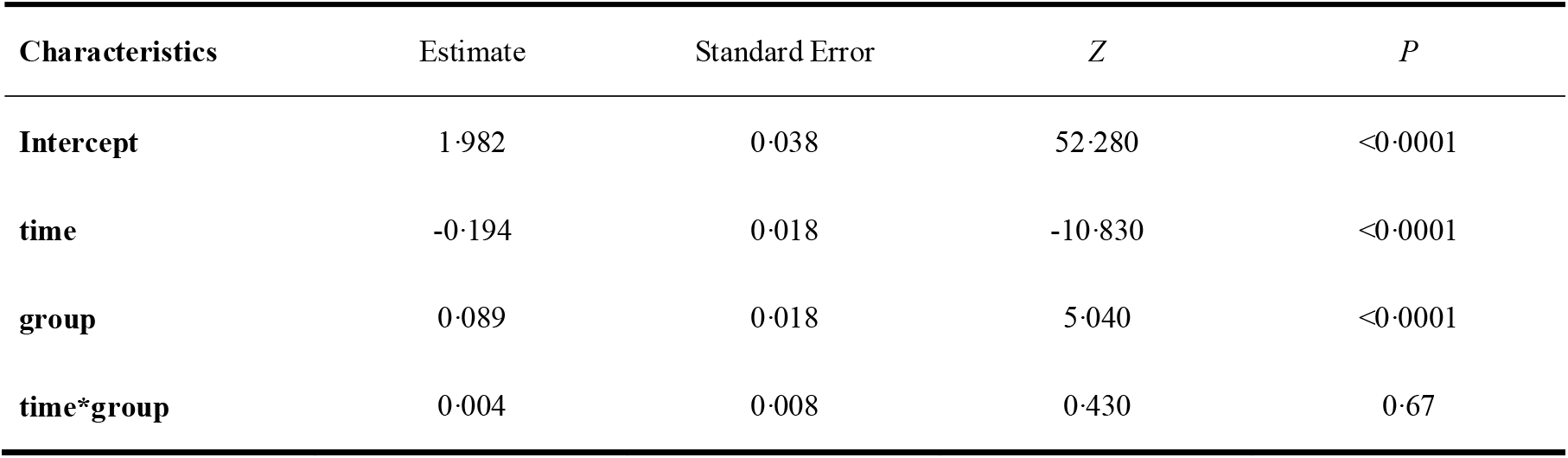
Kinetics of SARS-CoV-2 neutralizing antibody decay from day 28 to months 10 after the second dose vaccination by generalized estimating equations.

**Table 3:** The differences of SARS-CoV-2 neutralizing antibody titers between different follow-up point by paired.

| group | Time interval | Fold | Time interval | Fold |
| --- | --- | --- | --- | --- |
| 0-14d-10m | D<sub>28</sub>--M<sub>3</sub><sup>*</sup> | 2.12<sup>d</sup> | M<sub>3</sub>--M<sub>6</sub><sup>*</sup> | 1.52<sup>d</sup> |
|  | D<sub>28</sub>--M<sub>6</sub><sup>*</sup> | 3.22<sup>d</sup> | M<sub>3</sub>--M<sub>10</sub><sup>*</sup> | 2.29<sup>d</sup> |
|  | D<sub>28</sub>--M<sub>10</sub><sup>*</sup> | 4.85<sup>d</sup> | M<sub>6</sub>--M<sub>10</sub><sup>*</sup> | 1.50<sup>d</sup> |
|  | D<sub>28</sub>--D<sub>28</sub><sup>b#</sup> | 2.50<sup>i</sup> | M<sub>10</sub>--D<sub>28</sub><sup>b#</sup> | 12.16<sup>i</sup> |
| 0-21d-10m | D<sub>28</sub>--M<sub>3</sub><sup>*</sup> | 2.09<sup>d</sup> | M<sub>3</sub>--M<sub>6</sub><sup>*</sup> | 1.50<sup>d</sup> |
|  | D<sub>28</sub>--M<sub>6</sub><sup>*</sup> | 3.14<sup>d</sup> | M<sub>3</sub>--M<sub>10</sub><sup>*</sup> | 2.23<sup>d</sup> |
|  | D<sub>28</sub>--M<sub>10</sub><sup>*</sup> | 4.67<sup>d</sup> | M<sub>6</sub>--M<sub>10</sub><sup>*</sup> | 1.49<sup>d</sup> |
|  | D<sub>28</sub>--D<sub>28</sub><sup>b#</sup> | 2.06<sup>i</sup> | M<sub>10</sub>--D<sub>28</sub><sup>b#</sup> | 9.64<sup>i</sup> |
| 0-28d-10m | D<sub>28</sub>--M<sub>3</sub><sup>*</sup> | 2.03<sup>d</sup> | M<sub>3</sub>--M<sub>6</sub><sup>*</sup> | 1.52<sup>d</sup> |
|  | D<sub>28</sub>--M<sub>6</sub><sup>*</sup> | 3.09<sup>d</sup> | M<sub>3</sub>--M<sub>10</sub><sup>*</sup> | 2.21<sup>d</sup> |
|  | D<sub>28</sub>--M<sub>10</sub><sup>*</sup> | 4.49<sup>d</sup> | M<sub>6</sub>--M<sub>10</sub><sup>*</sup> | 1.45<sup>d</sup> |
|  | D<sub>28</sub>--D<sub>28</sub><sup>b#</sup> | 1.98<sup>i</sup> | M<sub>10</sub>--D<sub>28</sub><sup>b#</sup> | 8.91<sup>i</sup> |

D<sub>28</sub>: Day 28 after second dose; M<sub>3</sub>: Month 3 after second dose; M<sub>6</sub>: Month 6 after second dose; M<sub>10</sub>: Month 10 after second dose;
D<sub>28</sub><sup>b</sup>: Day 28 after booster dose
<sup>\*</sup> *P*-value of <0.008 (0.05/6) was considered significant
<sup>#</sup> *P*-value of <0.05 was considered significant
<sup>d</sup> Declining fold of neutralizing antibody GMT
<sup>i</sup> Increasing fold of neutralizing antibody GMT

**Figure 6:**
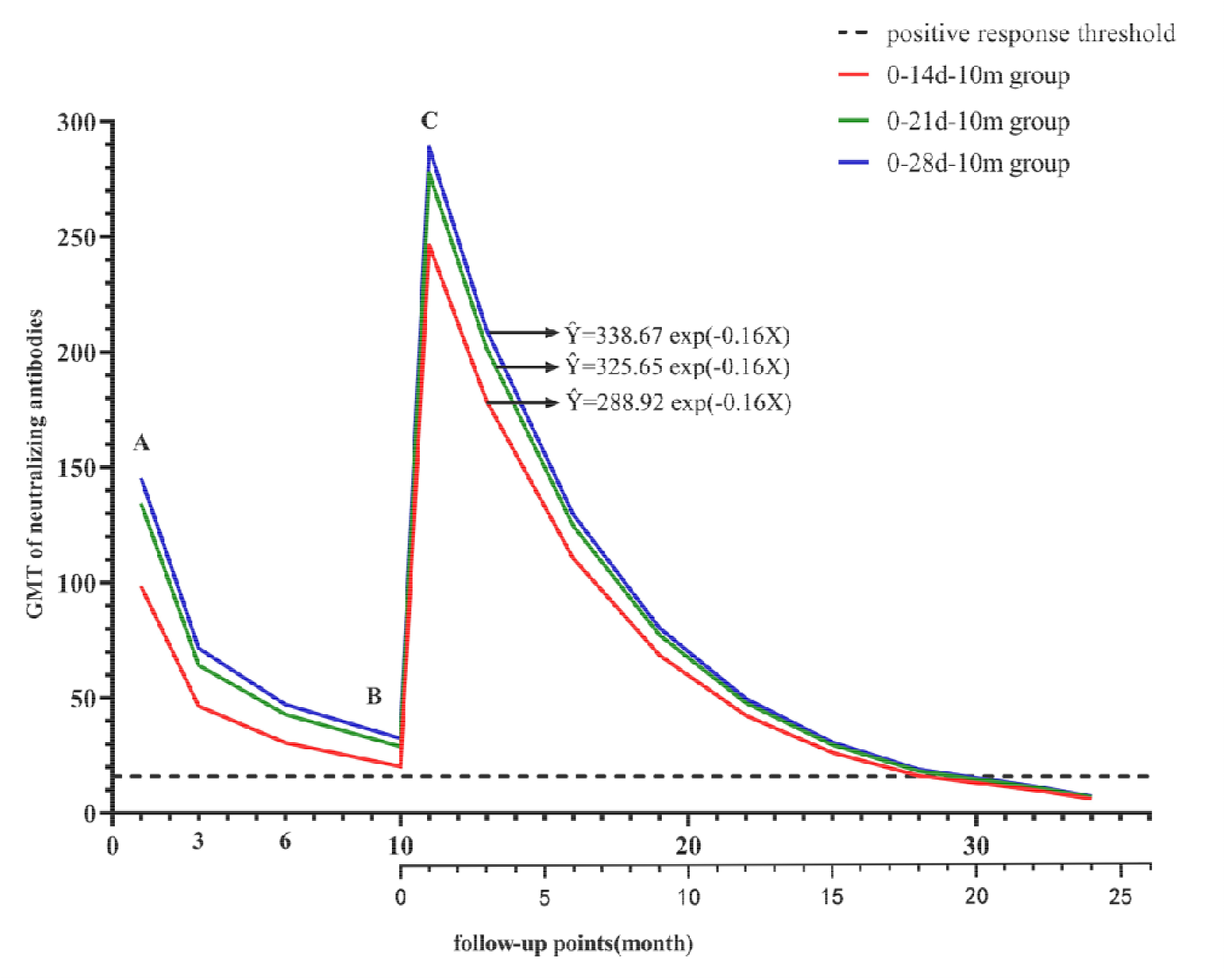
Kinetics of the GMT of SARS-CoV-2 neutralizing antibodies. **A:** Day 28 after the second dose; B: Month 10 after the second dose, booster dose; C: Day 28 after the booster dose. The curve from point A to point C is the actual observed value of follow-up, and the curve after point C is the predicted value by exponential curve model. The bottom axis is the time after booster dose vaccination. The dotted horizontal line represents the positive antibody response threshold.

The GMT of neutralizing antibodies from day 28 to month 10 of the second dose in 0-14d-10m group, 0-21d-10m group and 0-28d-10m group were used to construct decreased exponent curve model, respectively. The exponential curve equation was Ŷ=91.38 exp(−0.16X), Ŷ=127.10 exp(−0.16X) and Ŷ=138.38 exp(−0.16X) in three groups, respectively. The coefficient of determination *R* Squared was 0.85, 0.86 and 0.86 respectively. The antibody attenuation after booster dose was predicted according to the exponential curve model of antibody decay after two doses. The results of the three groups on the day 28 after booster dose were respectively substituted into the exponent model, time 28d (calculated according to 1 month) = 1, and the calculated parameter *α*values were 288.92, 325.65 and 338.67 respectively. So, the exponential curve equation for the 0-14d-10m group was Ŷ=288.92 exp(−0.16X), for the 0-21d-10m group was Ŷ=325.65 exp(−0.16X), and for the 0-28d-10m group was Ŷ=338.67 exp(−0.16X). It can be seen that the GMT of neutralizing antibodies may decrease by approximately 55% on 6 months post-booster and the predictive duration of booster-induced neutralizing antibody declining to the cutoff level of positive antibody response (GMT of 16) may be 18.08 months, 18.83 months and 19.08 months in three groups, respectively (Fig. 6; see appendix p 13-15).

### Predictors of SARS-CoV-2 neutralizing antibody durability within 10 months after the second dose

The results of Cox proportional hazards regression showed that the participants with age ≥40 (*HR*=1.883, 95%*CI*: 1.064∼3.333, *P*=0.03) were associated with a high risk of response loss (GMT<16). The participants who were in 0-28d-10m group (*HR*=0.353, 95%*CI*: 0.182∼0.683, *P*=0.0020), had an influenza vaccination history (*HR*=0.554, 95%*CI*: 0.336∼0.912, *P*=0.02) or were female (*HR*=0.299, 95%*CI*: 0.151∼0.593, *P*=0.0005) tended to maintain immune persistence during 10 months after the second dose (Fig. 7). The four factors above were used to construct a nomogram to predict neutralizing antibody decline probability during 10 months. The nomogram showed that the largest contributions of maintaining immune persistence were age and sex, followed by group and influenza vaccination history (Fig. 8). The C-index for the prediction nomogram was 0.70 for the training cohort (see appendix p 11).

**Figure 7:**
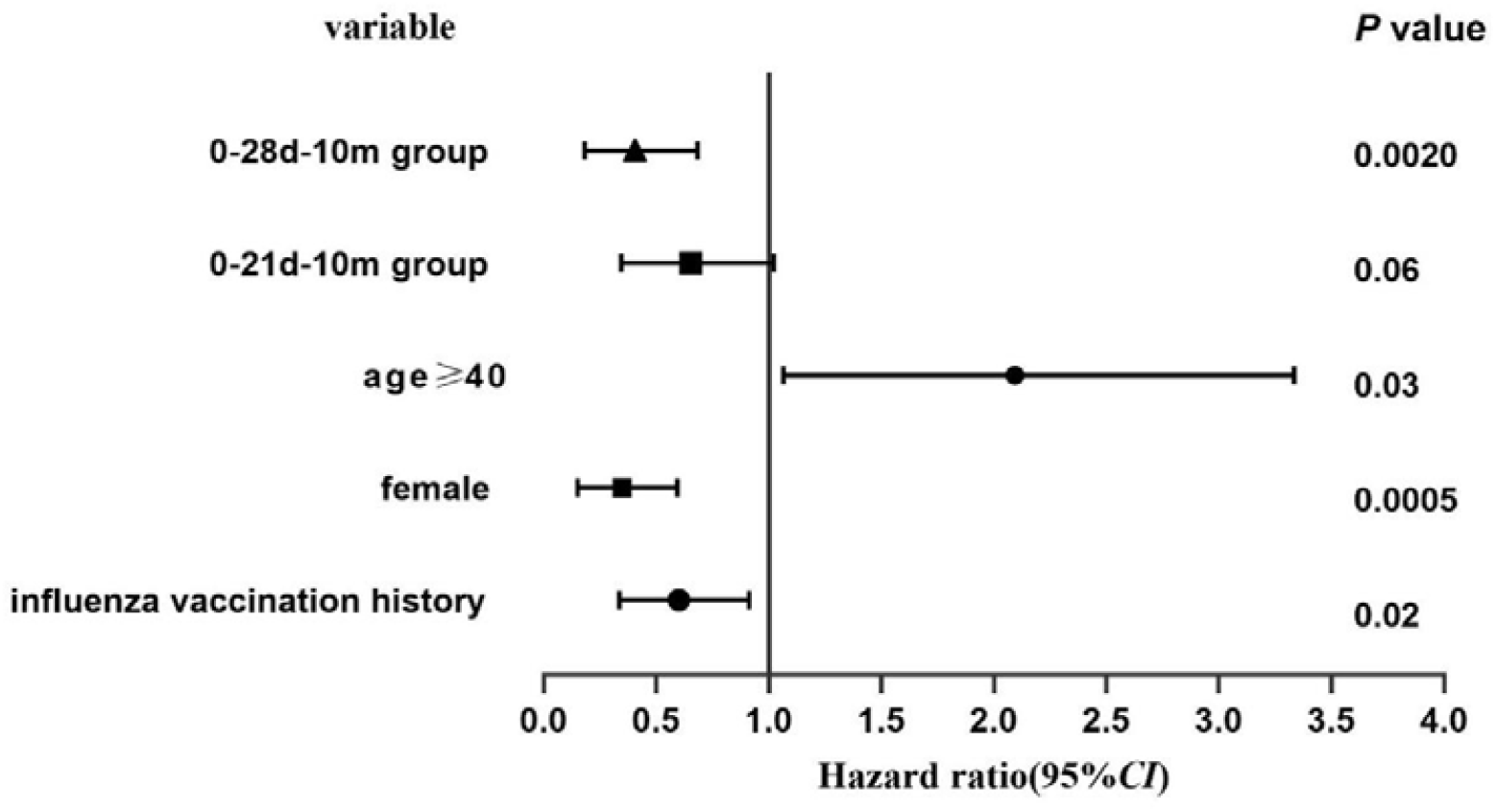
Influence factors of antibody loss by cox proportional hazard regression analysis.

**Figure 8:**
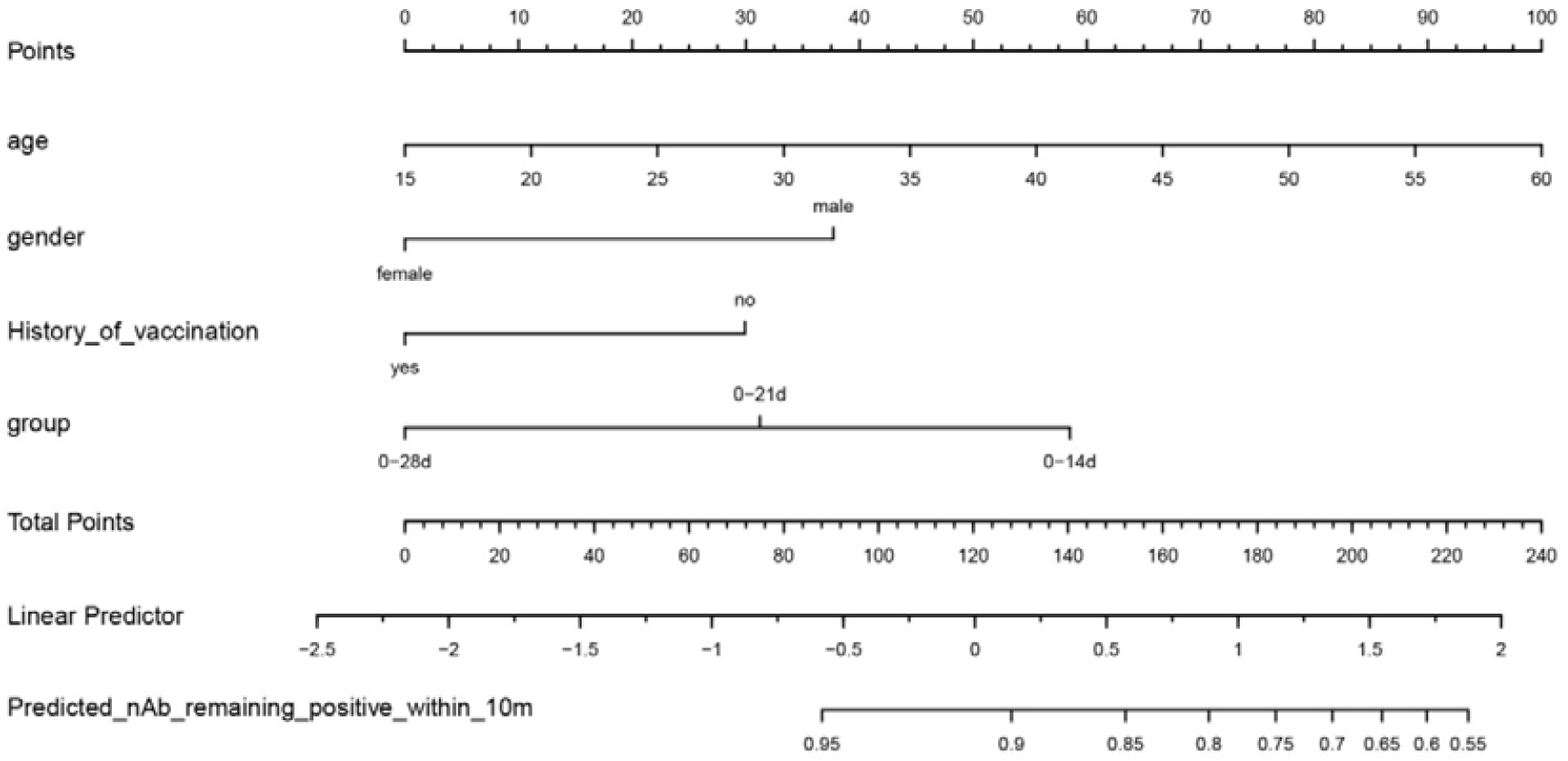
The predictors of the long-term immune persistence based on nomogram. The nomogram consists of graphical lines including predictors [age, sex, history of vaccination (The type of vaccine is influenza vaccine), difference groups], single score, total score, linear predictive value and event (Predicted of neutralizing antibody (nAb) remaining positive within 10 months). The scale is marked on the line segment corresponding to each predictor to represent the value range of the predictor, and the length of the line segment reflects the contribution of the predictor to the outcome event. The top single score in the picture shows the corresponding score of predictors under different values. The total score of all predictors is scored by single score. The corresponding linear prediction can be obtained. The lowest line represents the probability of SARS-CoV-2 neutralizing antibody remaining positive within 10 months.

### Safety

A total of 390 participants who complete the booster dose were included in safety analysis, the overall incidence of adverse reactions was 1.5% (2/130), 4.6% (6/130), and 3.1% (4/130) in the 0-14d-10m, 0-21d-10m and 0-28d-10m group after the booster dose, with no significant difference among the three groups (*P*=0.41). The most common reported reaction was injection-site pain, which occurred in 2 of 130 (1.5%), 3 of 130 (2.3%), and 1 of 130 (0.8%) participants in the three groups (*P*=0.87). Solicited adverse reactions were reported by 2 (1.5%) participants in the 0-14d-10m group, 4 (3.1%) participants in the 0-21d-10m group, and 3 (2.3%) participants in the 0-28d-10m group within 7 days after injection. All adverse reactions were mild after the booster injection. The vaccine showed a favorable safety profile from month 3 to 10 after second dose. None of the participants were infected SARS-CoV-2 during the trial period (Fig. 9; see appendix p 12).

**Figure 9:**
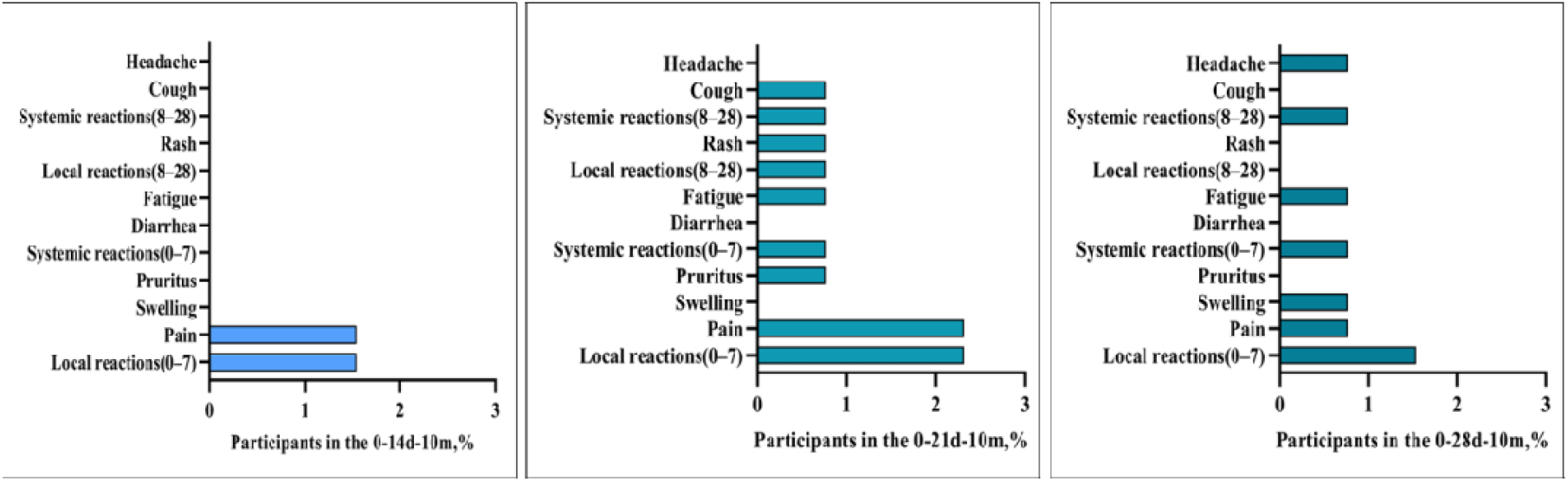
Incidence of solicited and unsolicited adverse reactions occurred within 28 days after the booster dose.

## Discussion

Recent studies reported the long-term immune persistence after the priming full-schedule vaccination and the safety and immunogenicity of booster dose with different technical platform COVID19 vaccines in different population [11-17, 21-23]. However, a complete picture of the kinetics of immune response among persons with different vaccination regimens of BBIBP-CorV is not yet available, and the safety and immunogenicity of homologous booster vaccination are yet to be thoroughly evaluated, especially in the high-risk occupational population. Based on our previous research, we assessed immune persistence of a priming two-dose BBIBP-CorV vaccine schedule, and found that the long-term persistent immunogenicity in 0-21d-10m and 0-28d-10m group was more satisfactory than that in 0-14d-10m group in public security officers and airport ground staff. Besides, maintaining long-term immune persistence was also associated with age<40, female, and history of influenza vaccination. A booster dose given at month 10 after the second dose led to a strong increase in neutralizing antibody which was not affected by the priming two-dose vaccination regimen, and the predictive duration of neutralizing antibody declining to the cutoff level of positive antibody response may be above 18 months in three groups.

To our knowledge, this research firstly reported the finding that a priming two-dose schedule of day 0-28 and day 0-21 could lead to longer persistence of neutralizing antibody compared to day 0-14 in the high-risk occupational population. Our initial study demonstrated that a priming vaccination regimen of day 0-21 and day 0-28 induced higher peak value of neutralizing antibodies at day 28 after the second dose [3]. Previous studies [25] demonstrated that the persistence of immune response was correlated with the peak antibody levels. That may account for the higher neutralizing antibody levels at each follow-up point from month 3 to month 10 after the second dose in 0-21d-10m and 0-28d-10m group.

Our results suggested that from day 28 to month 10 after the second dose, a decrease by 4.85-fold (GMT: 94.4-20.3), 4.67-fold (GMT: 134.4-28.8) and 4.49-fold (GMT: 145.5-32.4) was observed in the 0-14d-10m, 0-21d-10m and 0-28d-10m group, respectively. Other studies also evaluated the persistence of neutralizing antibodies induced by different inactivated vaccines. Zeng et al [13] found that neutralizing antibody titers induced by two doses (two schedules of day 0-14, day 0-28) CoronaVac vaccines (3 μg) both declined after 6 months to below the seropositive cutoff (GMT of 8). Another study [14] reported that the neutralizing antibodies induced by the two doses of BBIBP-CorV (3μg, 28 days apart) decreased by a factor of 2.94 (GMT: from 6.8 to 2.3) from month 2 to month 6. Similarly, one non-peer-reviewed study [16] in Indian found that the GMT of neutralizing antibodies decreased 8.24-fold from 28 days after a second BBV152 vaccination to 6 months later. In addition, neutralizing antibodies decreased over time after initial vaccination has been observed with mRNA vaccines and adenovirus-vectored vaccines et al, but to a lesser extent than those of inactivated vaccines [12, 15]. Overall, neutralizing antibody responses elicited by all kinds of COVID-19 vaccines after priming full-schedule vaccination, subsequently declining to varying degrees over time, which may be due to differences in vaccine technology platforms, study population or test methods et al.

Published data revealed that innate and adaptive immunity was decreased with age, particularly in vaccine responses [26]. Wang and colleagues [17] found that geometric mean concentrations of both inactivated vaccines (BBIBP-CorV and CoronaVac) decreased with age, especially in the 50-59 age group. A prospective longitudinal cohort study [18] also found that humoral response was substantially decreased 6 months after receipt of the second dose of the BNT162b2 vaccine, especially among persons 65 years of age or older. Consistent with previous studies, our results also found that the participants with age≥40 were associated with a high risk of response loss. Meanwhile, we found that persistence of neutralizing antibodies was better in women than in men, which was similar to the findings in other reports [17, 18]. It probably caused by immune function differences between the sexes. Mechanisms implicated in mediating sex-based differences in immune responses may include immunological, sex steroid hormones, genetic and epigenetic regulation, and microbiota differences et al [27].

We also found that history of influenza vaccination had a positive impact on the immune persistence of the SARS-CoV-2 vaccine [The median time from the influenza vaccination to the first dose of BBIBP-CorV vaccine was 80.74 days, interquartile range (IQR) 71.70-90.74]. A real-world study [28] revealed that influenza/pneumococcal vaccination seems to have a substantial impact on the neutralization response to BNT162b2 mRNA vaccination (The median time from the influenza and pneumococcal vaccination to the second dose of SARS-CoV-2 vaccine was 102 days). It is speculated that influenza vaccine may affect the persistence of the COVID-19 vaccine by affecting its immunogenicity. It may be related to the non-specific effects of vaccines [29].

Several studies reported that the homologous booster dose with inactivated vaccines led to a strangely rebound in neutralizing immune response against SARS-CoV-2. A non-peer-reviewed study [21] found the GMT of neutralizing antibodies was improved 1.04-3.87-fold on day 30 after a booster dose of BBIBP-CorV vaccine compared to the 28 days after the second dose. Similarly, one study [14] of 353 healthy adult participants who administered with a two-shot regiment (28 days apart) BBIBP-CorV vaccine (month 1) and a booster dose 7 months later, the GMT increased 4.5 times, compared to GMT of month 2. Another non-peer-reviewed study [30] reported that a booster vaccination at 8 to 9 months after priming two-dose BBIBP-CorV vaccine led to a 6.1-fold increase in the neutralization GMT against the wild-type strain. Besides, some studies also revealed that a heterologous boost following prime vaccination could also elicit remarkably protection against SARS-CoV-2 [21, 23]. Consistent with previous studies, our research indicated that on 28 days after the homologous booster dose, GMT of neutralizing antibodies were increased by 8.91-12.16-fold compared to the pre-boosting levels and 1.98-2.50 fold compared to the 28 days after the second dose levels. Overall, the booster dose could significantly reverse the decrease in neutralizing antibodies after the second dose, which may be explained by that the priming two-dose vaccination could induce efficient T and B memory cells and booster vaccination could significantly recall and enhance antibody responses [8, 9, 16].

Notably, our results indicated that a booster dose given at month 10 after the second dose produced similar immunogenicity (246.2, 277.5 and 288.6) among the three groups, which was not affected by the priming two-dose vaccination regimen. It means that booster dose could reverse the lower GMT of neutralizing antibodies of 14 days intervals at some extent. Meanwhile the results also highlighted the essential for booster vaccination to top-up the immune response. Of course, these results should be confirmed by large-scale clinical studies.

Furthermore, using the exponent curve model, we predicted that the GMT of neutralizing antibodies may decrease by approximately 55% at 6 months post-booster and the predictive duration of neutralizing antibodies declining to the cutoff level of positive antibody response may be above 18 months in three groups. One non-peer-reviewed study [31] in China reported that the GMT of neutralization antibody against an ancestral SARS-CoV-2 viral strain (Wuhan-Hu-1) drastically decreased by approximately 85% at 6 months after the booster dose of inactivated vaccines. Our model prediction results showed a satisfactory long-term durability of neutralizing antibodies in high-risk occupational population, and data will be collected as verified the prediction results in the future.

This study has some limitations. Firstly, neutralization tests in vitro against emerging Omicron variant were not assessed in our study. Several studies have shown vaccine-induced immune protection might more likely be escaped by Omicron compared to prototypes and other VOCs, while a booster dose improved neutralization against Omicron [30, 32, 33], and we will further explore. Secondly, we only reported persistence of a two-dose schedule data and immunogenicity and safety of a booster dose data for the high-risk occupational population aged 18 to 59 years. Although the population aged 60 years and older was not evaluated in this study, our study still suggested that older people (age≥40) have faster neutralizing antibody decay. Thirdly, the C-index of our nomogram was not satisfactory enough, however, it could reflect the predictors of antibody decay in some extent. Finally, a relatively high rate of loss to follow-up is unavoidable during the follow-up. However, there were no differences in the rates of loss to follow-up among the three groups.

## Conclusions

In conclusion, the priming two-dose BBIBP-CorV vaccine with 0-28 days and 0-21 days schedule could lead a longer persistence of neutralizing antibody than 0-14 days schedule. Maintaining long-term immune persistence was also associated with age<40, female, and history of influenza vaccination. Regardless of priming two-doses vaccination regimens, a homologous booster dose led to a strong rebound in neutralizing antibody and might elicit satisfactory persistent immunity.

## Supporting information

protocol 1

protocol 2

## Data Availability

In this study, the full protocol and the datasets are available, following manuscript publication, upon request from the corresponding author (Professor Suping Wang), following the provision of ethics approval.

## Abbreviations

SARS-CoV-2: Severe acute respiratory syndrome coronavirus 2;
COVID-19: Coronavirus disease 2019;
GMT: Geometric mean titer;
CI: Confdence interval; BMI: Body mass index;
RT-PCR: Reverse transcriptase-polymerase chain reaction;
WHO: World Health Organization.

## Acknowledgments

This research was supported by the COVID-19 Project of Shanxi Provincial Finance, and the Project of Shanxi Provincial Key Laboratory for major infectious disease response. Thanks for gratefully all participants taking part in this research. We gratefully acknowledge the contribution from our colleagues and students, and staff members of the Shanxi Provincial Center for Disease Control and Outpatient Department of Shanxi Aviation Industry Group Co.LTD.

## Authors’ Contributors

Tian Yao: conceptualization, conceived and developed the overall study design, reviewed and interpreted data, investigated and performed experiments, and drafted the manuscript. Xiaohong Zhang, Shengcai Mu: conceptualization, conceived and designed the study, analyzed the data, reviewed and interpreted data, investigated and performed experiments. Yana Guo, Xiuyang Xu: analyzed the data, reviewed and interpreted data, investigated and performed experiments. Junfeng Huo, Zhiyun Wei, Ling Liu, Xiaoqing Li, Hong Li, Rongqin Xing: investigated and performed experiments. Yongliang Feng & Jing Chen & Lizhong Feng & Suping Wang: conceptualization, conceived and designed the study, reviewed and interpreted data, writing-review & editing.

## Funding

The study was supported by the COVID-19 Project of Shanxi Provincial Finance, the Project of Shanxi Provincial Key Laboratory for major infectious disease response, and the COVID-19 Scientific Research Project of Health Commission of Shanxi Provincial.

## Declarations

### Ethics approval and consent to participate

The study was approved by the Ethics Committee of Shanxi Provincial Center for Disease Control (SXCDCIRBPJ2020056001) and Prevention and the Chinese Clinical Trial Registry (ChiCTR2100041705, ChiCTR2100041706).

### Consent for publication

Not applicable.

### Competing interests

All authors declare that they have no competing interests.

## Notes

### Competing Interest Statement

The authors have declared no competing interest.

### Clinical Trial

ChiCTR2100041705，ChiCTR2100041706

### Author Declarations

The study was approved by the Ethics Committee of Shanxi Provincial Center for Disease Control (SXCDCIRBPJ2020056001) and Prevention

