## supplementary appendix for "Long-term immune persistence induced by two-dose BBIBP-CorV vaccine with different intervals, and immunogenicity and safety of a homologous booster dose in high-risk occupational population Secondary Study Based on a Randomized Clinical Trial"

**Table of Contents**

**Supplemental of Missing value fill…………………………………………………..2**

Table 1. Details about observed data and complete case data…………………. ……..2

Figure S1: Process on how missing measurements of neutralizing antibody titers were imputed………………………………………………………………………………...3

**Supplemental Results of……………………………………………………………...4**

Table S2. Demographic and Behavioral Characteristics of High-risk Occupational Population at Day 28 after the Second Dose………………………………….……….4

Table S3. Demographic and Behavioral Characteristics of High-risk Occupational Population at Month 3 after the Second Dose…………………………………………5

Table S4. Demographic and Behavioral Characteristics of High-risk Occupational Population at Month 6 after the Second Dose…………………………………………6

Table S5. Demographic and Behavioral Characteristics of High-risk Occupational Population at Month 10 after the Second Dose………………………………………..7

Table S6. Demographic and Behavioral Characteristics of High-risk Occupational Population who Received Booster Dose of Vaccine and no Vaccination……………..8

**Supplemental Results of……………………………………………………………...9**

Table S7. Positive rates of SARS‑CoV‑2 neutralizing antibody at each follow-up point in three groups…………………………………………………………………...9

Table S8. Constituent ratio of SARS‑CoV‑2 neutralizing antibody at each follow-up point in three groups………………………………………………………………….10

**Supplemental Results of nomogram……………………………………………….11**

Figure S2. The calibration curves for the nomogram…………………………….......11

Figure S3. Calibration curve for internal verification of the prediction tool……...…11

Supplemental Results of safety…………………………………………………….12

Table S9. Summary of solicited and unsolicited adverse reactions occurred within 28 days after the third dose……………………………………………………………...12

**Supplemental Methods……………………...……………………………………...13**

Basic principles…………………………………...………………………………….13

Fitting process………………………………………………………………………..13

Table S1. Details about observed data and complete case data

|  | **No. of participants** | | | | | | | |
| --- | --- | --- | --- | --- | --- | --- | --- | --- |
|  | **0-14d-10m group** | |  | **0-21d-10m group** | |  | **0-28d-10m group** | |
|  | **Responders** | **Non responders** |  | **Responders** | **Non responders** |  | **Responders** | **Non responders** |
| **Observed at 28d** | 256 | 0 |  | 247 | 0 |  | 241 | 0 |
| **Month 3** |  |  |  |  |  |  |  |  |
| Observed data | 233 | 9 |  | 227 | 2 |  | 221 | 3 |
| Imputed Responses/nonresponses^a^ | 4 | 0 |  | 6 | 0 |  | 7 | 0 |
| Complete case data^b^ | 237 | 9 |  | 233 | 2 |  | 228 | 3 |
| **Month 6** |  |  |  |  |  |  |  |  |
| Observed data | 195 | 6 |  | 189 | 6 |  | 167 | 2 |
| Imputed Responses/nonresponses^a^ | 9 | 9 |  | 17 | 2 |  | 22 | 3 |
| Complete case data^b^ | 204 | 15 |  | 206 | 8 |  | 189 | 5 |
| **Month 10** |  |  |  |  |  |  |  |  |
| Observed data | 124 | 25 |  | 130 | 15 |  | 123 | 11 |
| Imputed Responses/nonresponses^a^ | 8 | 15 |  | 17 | 8 |  | 18 | 5 |
| Complete case data^b^ | 132 | 40 |  | 147 | 23 |  | 141 | 16 |

^a^ We followed up participants who were antibody positive at day 28 after priming two-dose vaccine. During the follow-up, if the neutralizing antibody response of participants was negative, they will no longer be followed up, and the antibody response at each time point thereafter was negative by default. When the previous observed titer was positive and still positive in the next follow-up, we imputed the intermediate missing value as a positive response and the antibody titers were filled according to the next follow-up. If neutralizing antibody was positive at the previous follow-up and negative at the next follow-up, the intermediate missing value would not be filled.

^b^ The imputed nonresponses and responses and the observed neutralizing antibody levels were together referred to as complete case data.

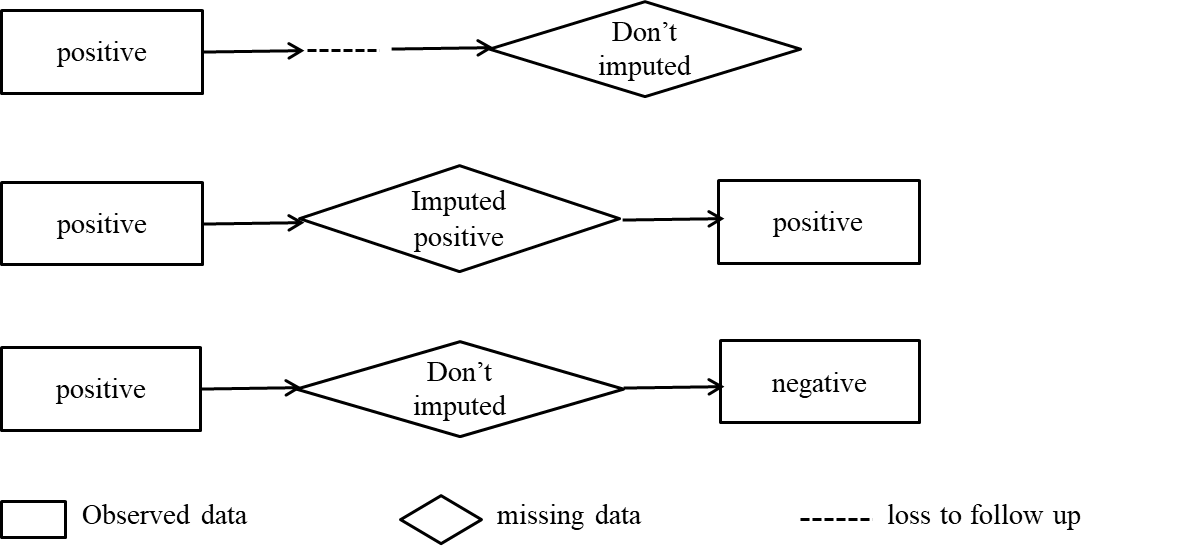

Figure S1: Process on how missing measurements of neutralizing antibody titers were imputed.

Table S2. Demographic and Behavioral Characteristics of High-risk Occupational Population at Day 28 after the Second Dose

| **Characteristics** | **Total** | **0-14 group** | **0-21 group** | **0-28 group** | ***P*** |
| --- | --- | --- | --- | --- | --- |
|  | **n=744** | **n=256** | **n=247** | **n=241** |  |
| gender |  |  |  |  | 0.49 |
| Male | 538(72.3%) | 179(69.9%) | 179(72.5%) | 180(74.7%) |  |
| Female | 206(27.7%) | 77(30.1%) | 68(27.5%) | 61(25.3%) |  |
| Age(years) |  |  |  |  | 0.41 |
| <40 | 427(57.4%) | 139(54.3%) | 143(57.9%) | 145(60.2%) |  |
| ≥40 | 317(42.6%) | 117(45.7%) | 104(42.1%) | 96(39.8%) |  |
| Education level |  |  |  |  | 0.22 |
| Junior high school or lower | 70(9.4%) | 31(12.1%) | 23(9.3%) | 16(6.6%) |  |
| Senior high school | 33(4.4%) | 10(3.9%) | 9(3.6%) | 14(5.8%) |  |
| College or higher | 641(86.2%) | 215(84.0%) | 215(87.1%) | 211(87.6%) |  |
| Maritial status |  |  |  |  | 0.16 |
| Married | 573(77.0%) | 205(80.1%) | 179(72.5%) | 189(78.4%) |  |
| Unmarried | 150(20.2%) | 45(17.6%) | 62(25.1%) | 43(17.8%) |  |
| Divorced or widowed | 21(2.8%) | 6(2.3%) | 6(2.4%) | 9(3.8%) |  |
| Ethnicity |  |  |  |  | 0.48 |
| Han ethnicity | 733(98.5%) | 254(99.2%) | 242(98.0%) | 237(98.3%) |  |
| Other | 11(1.5%) | 2(0.8%) | 5(2.0%) | 4(1.7%) |  |
| BMI (kg/m^2^) |  |  |  |  | 0.64 |
| <24 | 327(43.9%) | 115(44.9%) | 112(45.3%) | 100(41.5%) |  |
| ≥24 | 417(56.1%) | 141(55.1%) | 135(54.7%) | 141(58.5%) |  |
| Influenza vaccination history |  |  |  |  | 0.93 |
| No | 497(66.8%) | 169(66.0%) | 167(67.6%) | 161(66.8%) |  |
| Yes | 247(33.2%) | 87(34.0%) | 80(32.4%) | 80(33.2%) |  |

Results expressed as n (%), BMI=body-mass index.

Table S3. Demographic and Behavioral Characteristics of High-risk Occupational Population at Month 3 after the Second Dose

| **Characteristics** | **Total** | **0-14 group** | **0-21 group** | **0-28 group** | ***P*** |
| --- | --- | --- | --- | --- | --- |
|  | **n=712** | **n=246** | **n=235** | **n=231** |  |
| gender |  |  |  |  | 0.54 |
| Male | 509(71.5%) | 171(69.5%) | 167(71.1%) | 171(74.0%) |  |
| Female | 203(28.5%) | 75(30.5%) | 68(28.9%) | 60(26.0%) |  |
| Age(years) |  |  |  |  | 0.37 |
| <40 | 407(57.2%) | 132(53.7%) | 137(58.3%) | 138(59.7%) |  |
| ≥40 | 305(43.8%) | 114(46.3%) | 98(41.7%) | 93(40.3%) |  |
| Education level |  |  |  |  | 0.28 |
| Junior high school or lower | 69(9.7%) | 30(12.2%) | 23(9.8%) | 16(6.9%) |  |
| Senior high school | 33(4.6%) | 10(4.1%) | 9(3.8%) | 14(6.1%) |  |
| College or higher | 610(85.7%) | 206(83.7%) | 203(86.4%) | 201(87.0%) |  |
| Maritial status |  |  |  |  | 0.08 |
| Married | 550(77.3%) | 198(80.5%) | 169(71.9%) | 183(79.2%) |  |
| Unmarried | 142(19.9%) | 43(17.5%) | 60(25.5%) | 39(16.9%) |  |
| Divorced or widowed | 20(2.8%) | 5(2.0%) | 6(2.6%) | 9(3.9%) |  |
| Ethnicity |  |  |  |  | 0.48 |
| Han ethnicity | 701(98.5%) | 244(99.2%) | 230(97.9%) | 227(98.3%) |  |
| Other | 11(1.5%) | 2(0.8%) | 5(2.1%) | 4(1.7%) |  |
| BMI (kg/m^2^) |  |  |  |  | 0.41 |
| <24 | 318(44.7%) | 113(45.9%) | 110(46.8%) | 95(41.1%) |  |
| ≥24 | 394(55.3%) | 133(54.1%) | 125(53.2%) | 136(58.9%) |  |
| Influenza vaccination history |  |  |  |  | 0.99 |
| No | 470(66.0%) | 162(65.9%) | 156(66.4%) | 152(65.8%) |  |
| Yes | 242(34.0%) | 84(34.1%) | 79(33.6%) | 79(34.2%) |  |

Results expressed as n (%), BMI=body-mass index.

Table S4. Demographic and Behavioral Characteristics of High-risk Occupational Population at Month 6 after the Second Dose

| **Characteristics** | **Total** | **0-14 group** | **0-21 group** | **0-28 group** | ***P*** |
| --- | --- | --- | --- | --- | --- |
|  | **n=627** | **n=219** | **n=214** | **n=194** |  |
| gender |  |  |  |  | 0.45 |
| Male | 444(70.8%) | 152(69.4%) | 148(69.2%) | 144(74.2%) |  |
| Female | 183(29.2%) | 67(30.6%) | 66(30.8%) | 50(25.8%) |  |
| Age(years) |  |  |  |  | 0.37 |
| <40 | 370(59.0%) | 122(55.7%) | 127(59.3%) | 121(62.4%) |  |
| ≥40 | 257(41.0%) | 97(44.3%) | 87(40.7%) | 73(37.6%) |  |
| Education level |  |  |  |  | 0.29 |
| Junior high school or lower | 64(10.2%) | 28(12.8%) | 21(9.8%) | 15(7.7%) |  |
| Senior high school | 30(4.8%) | 9(4.1%) | 8(3.7%) | 13(6.7%) |  |
| College or higher | 533(85.0%) | 182(83.1%) | 185(86.5%) | 166(85.6%) |  |
| Maritial status |  |  |  |  | 0.24 |
| Married | 479(76.4%) | 175(79.9%) | 152(71.0%) | 152(78.3%) |  |
| Unmarried | 132(21.0%) | 39(17.8%) | 56(26.2%) | 37(19.1%) |  |
| Divorced or widowed | 16(2.6%) | 5(2.3%) | 6(2.8%) | 5(2.6%) |  |
| Ethnicity |  |  |  |  | 0.71 |
| Han ethnicity | 618(98.6%) | 217(99.1%) | 210(98.1%) | 191(98.5%) |  |
| Other | 9(1.4%) | 2(0.9%) | 4(1.9%) | 3(1.5%) |  |
| BMI (kg/m^2^) |  |  |  |  | 0.54 |
| <24 | 281(44.8%) | 99(45.2%) | 101(47.2%) | 81(41.7%) |  |
| ≥24 | 346(55.2%) | 120(54.8%) | 113(52.8%) | 113(58.3%) |  |
| Influenza vaccination history |  |  |  |  | 0.93 |
| No | 401(64.0%) | 139(63.5%) | 139(64.9%) | 123(63.4%) |  |
| Yes | 226(36.0%) | 80(36.5%) | 75(35.1%) | 71(36.6%) |  |

Results expressed as n (%), BMI=body-mass index.

Table S5. Demographic and Behavioral Characteristics of High-risk Occupational Population at Month 10 after the Second Dose

| **Characteristics** | **Total** | **0-14 group** | **0-21 group** | **0-28 group** | ***P*** |
| --- | --- | --- | --- | --- | --- |
|  | **n=499** | **n=172** | **n=170** | **n=157** |  |
| gender |  |  |  |  | 0.25 |
| Male | 340(68.1%) | 114(66.3%) | 111(65.3%) | 115(73.2%) |  |
| Female | 159(31.9%) | 58(33.7%) | 59(34.7%) | 42(26.8%) |  |
| Age(years) |  |  |  |  | 0.05 |
| <40 | 316(63.3%) | 97(56.4%) | 111(65.3%) | 108(68.8%) |  |
| ≥40 | 183(36.7%) | 75(43.6%) | 59(34.7%) | 49(31.2%) |  |
| Education level |  |  |  |  | 0.20 |
| Junior high school or lower | 56(11.2%) | 25(14.5%) | 19(11.2%) | 12(7.6%) |  |
| Senior high school | 29(5.8%) | 8(4.7%) | 8(4.7%) | 13(8.3%) |  |
| College or higher | 414(83.0%) | 139(80.8%) | 143(84.1%) | 132(84.1%) |  |
| Maritial status |  |  |  |  | 0.31 |
| Married | 375(75.2%) | 137(79.7%) | 119(70.0%) | 119(75.8%) |  |
| Unmarried | 110(22.0%) | 30(17.4%) | 46(27.1%) | 34(21.7%) |  |
| Divorced or widowed | 14(2.8%) | 5(2.9%) | 5(2.9%) | 4(2.5%) |  |
| Ethnicity |  |  |  |  | 0.83 |
| Han ethnicity | 491(98.4%) | 170(98.8%) | 167(98.2%) | 154(98.1%) |  |
| Other | 8(1.6%) | 2(1.2%) | 3(1.8%) | 3(1.9%) |  |
| BMI (kg/m^2^) |  |  |  |  | 0.84 |
| <24 | 229(45.9%) | 82(47.7%) | 77(45.3%) | 70(44.6%) |  |
| ≥24 | 270(54.1%) | 90(52.3%) | 93(54.7%) | 87(55.4%) |  |
| Influenza vaccination history |  |  |  |  | 0.94 |
| No | 280(56.1%) | 95(55.2%) | 97(57.1%) | 88(56.1%) |  |
| Yes | 219(43.9%) | 77(44.8%) | 73(42.9%) | 69(43.9%) |  |

Results expressed as n (%), BMI=body-mass index.

Table S6. Demographic and Behavioral Characteristics of High-risk Occupational Population who Received Booster Dose of Vaccine and no Vaccination

| **Characteristics** | **Total** | **unvaccinated group** | **vaccinated group** | ***P*** |
| --- | --- | --- | --- | --- |
|  | **n=809** | **n=419** | **n=390** |  |
| gender |  |  |  | 0.57 |
| Male | 592(73.2%) | 303(72.3%) | 289 (74.1%) |  |
| Female | 217(26.8%) | 116(27.7%) | 101(25.9%) |  |
| Age(years) |  |  |  | 0.13 |
| <40 | 463(57.2%) | 229(54.7%) | 234(60.0%) |  |
| ≥40 | 346(42.8%) | 190(45.3%) | 156(40.0%) |  |
| Education level |  |  |  | <0.001 |
| Junior high school or lower | 74(9.1%) | 28(6.7%) | 46(11.8%) |  |
| Senior high school | 37(4.6%) | 11(2.6%) | 26(6.7%) |  |
| College or higher | 698(86.3%) | 380(90.7%) | 318(81.5%) |  |
| Maritial status |  |  |  | 0.58 |
| Married | 621(76.8%) | 326(77.8%) | 295(75.6%) |  |
| Unmarried | 165(20.4%) | 80(19.1%) | 85(21.8%) |  |
| Divorced or widowed | 23(2.8%) | 13(3.1%) | 10(2.6%) |  |
| Ethnicity |  |  |  | 0.25 |
| Han ethnicity | 797(98.5%) | 415(99.1%) | 382(97.9%) |  |
| Other | 12(1.5%) | 4(0.9%) | 8(2.1%) |  |
| BMI (kg/m^2^) |  |  |  | 0.20 |
| <24 | 352(43.5%) | 173(41.3%) | 179(45.9%) |  |
| ≥24 | 457(56.5%) | 246(58.7%) | 211(54.1%) |  |
| Influenza vaccination history |  |  |  | 0.72 |
| No | 549(67.9%) | 282(67.3%) | 267(68.5%) |  |
| Yes | 260(32.1%) | 137(32.7%) | 123(31.5%) |  |

Results expressed as n (%), BMI=body-mass index.

Table S7. Positive rates of SARS‑CoV‑2 neutralizing antibody at each follow-up point in three groups

| **Time of assessment** | **Complete case analysis（%）** | | |
| --- | --- | --- | --- |
|  | **0–14d-10m group** | **0–21d-10m group** | **0–28d-10m group** |
| Day 28 after second dose |  |  |  |
| GMT (95%*CI*) | 98.4 (88.4–108.4)^a^ | 134.4(123.1-145.7)^b^ | 145.5(131.3-159.6)^b^ |
| GMT ≥ 16 | 256/256 (100.0%) | 247/247 (100.0%) | 241/241 (100.0%) |
| GMT ≥ 32 | 222/256 (86.7%)^a^ | 238/247 (96.4%)^b^ | 231/241 (95.9%)^b^ |
| GMT ≥ 64 | 156/256 (60.9%)^a^ | 209/247 (84.6%)^b^ | 195/241 (80.9%)^b^ |
| GMT ≥ 128 | 81/256 (31.6%)^a^ | 138/247 (55.9%)^b^ | 124/241 (51.5%)^b^ |
| GMT ≥ 256 | 23/256 (9.0%)^a^ | 37/247 (15.0%)^b^ | 42/241 (17.4%)^b^ |
| Month 3 after second dose |  |  |  |
| GMT (95%*CI*) | 46.4(41.4-52.0)^a^ | 64.2(58.8-70.1)^b^ | 71.6(65.7-78.0)^b^ |
| GMT ≥ 16 | 237/246(96.3%) | 233/235(99.2%) | 228/231(98.7%) |
| GMT ≥ 32 | 194/246 (78.9%)^a^ | 210/235(89.4%)^b^ | 215/231(93.1%)^b^ |
| GMT ≥ 64 | 110/246 (44.7%)^a^ | 140/235(59.6%)^b^ | 150/231(64.9%)^b^ |
| GMT ≥ 128 | 28/246(11.4%)^a^ | 44/235(18.7%)^b^ | 62/231(26.8%)^c^ |
| GMT ≥ 256 | 0/246(0.0%)^a^ | 4/235(1.7%) | 6/231(2.6%)^b^ |
| Month 6 after second dose |  |  |  |
| GMT (95%*CI*) | 30.5(27.0-34.5)^a^ | 42.8(39.2-46.7)^b^ | 47.1 (42.2-52.6)^b^ |
| GMT ≥ 16 | 204/219(93.2%) | 206/214(96.3%) | 189/194(97.4%) |
| GMT ≥ 32 | 139/219(63.5%)^a^ | 158/214(73.8%)^b^ | 156/194(80.4%)^b^ |
| GMT ≥ 64 | 41/219(18.7%)^a^ | 75/214(35.1%)^b^ | 84/194(43.3%)^b^ |
| GMT ≥ 128 | 5/219(2.3%)^a^ | 11/214(5.1%) | 13/194(6.7%)^b^ |
| GMT ≥ 256 | 0/219(0.0%) | 0/214(0.0%) | 0/194(0.0%) |
| Month 10 after second dose |  |  |  |
| GMT (95%*CI*) | 20.3(17.4-23.6)^a^ | 28.8 (25.5-32.6)^b^ | 32.4 (28.7-36.6)^b^ |
| GMT ≥ 16 | 132/172(76.7%)^a^ | 147/170(86.5%)^b^ | 141/157(89.8%)^b^ |
| GMT ≥ 32 | 75/172(43.6%)^a^ | 101/170(59.4%)^b^ | 93/157(59.2%)^b^ |
| GMT ≥ 64 | 13/172(7.6%)^a^ | 23/170(13.5%)^a^ | 35/157(22.3%)^b^ |
| GMT ≥ 128 | 0/172(0.0%) | 0/170(0.0%) | 2/157(1.3%) |
| GMT ≥ 256 | 0/172(0.0%) | 0/170(0.0%) | 0/157(0.0%) |
| Day 28 after booster dose |  |  |  |
| GMT (95%*CI*) | 246.2(222.6-269.7) | 277.5(248.9-306.0) | 288.6(256.9-320.3) |
| GMT ≥ 16 | 122/122(100.0%) | 120/120(100.0%) | 112/112(100.0%) |
| GMT ≥ 32 | 122/122(100.0%) | 120/120(100.0%) | 112/112(100.0%) |
| GMT ≥ 64 | 120/122(98.4%) | 119/120(99.2%) | 112/112(100.0%) |
| GMT ≥ 128 | 102/122(83.6%) | 109/120(90.8%) | 102/112(91.1%) |
| GMT ≥ 256 | 59/122(48.4%) | 68/120(56.7%) | 63/112(56.3%) |

Results expressed as n/N (%).

GMT: Geometric mean titer; *CI*: Confidence interval.

^a,b,c^ There was significant difference with the different letters.

Table S8. Constituent ratio of SARS‑CoV‑2 neutralizing antibody at each follow-up point in three groups

| **Time of assessment** | **Constituent ratio (%)** | | |
| --- | --- | --- | --- |
|  | **0-14d-10m** | **0-21d-10m** | **0-28d-10m** |
| day 28 after second dose |  |  |  |
| 16-31 | 13.3%(34/256) | 3.6%(9/247) | 4.2%(10/241) |
| 32-63 | 25.8%(66/256) | 11.7%(29/247) | 14.9%(36/241) |
| 64-127 | 29.3%(75/256) | 28.8%(71/247) | 29.5% (71/241) |
| 128-255 | 22.6% (58/256) | 40.9% (101/247) | 34.0% (82/241) |
| ≥256 | 9.0% (23/256) | 15.0% (37/247) | 17.4% (42/241) |
| Month 3 after second dose |  |  |  |
| 16-31 | 18.2% (43/237) | 9.9% (23/233) | 5.7% (13/228) |
| 32-63 | 35.4% (84/237) | 30.0% (70/233) | 28.5% (65/228) |
| 64-127 | 34.6% (82/237) | 41.2% (96/233) | 38.6% (88/228) |
| 128-255 | 11.8% (28/237) | 17.2% (40/233) | 24.6% (56/228) |
| ≥256 | 0.0% (0/237) | 1.7% (4/233) | 2.6% (6/228) |
| Month 6 after second dose |  |  |  |
| 16-31 | 31.9% (65/204) | 23.3% (48/206) | 17.4% (33/189) |
| 32-63 | 48.0% (98/204) | 40.3% (83/206) | 38.1% (72/189) |
| 64-127 | 17.7% (36/204) | 31.1% (64/206) | 37.6% (71/189) |
| 128-255 | 2. 4% (5/204) | 5.3% (11/206) | 6.9% (13/189) |
| ≥256 | 0.0% (0/204) | 0.0% (0/206) | 0.0% (0/189) |
| Month 10 after second dose |  |  |  |
| 16-31 | 43.2% (57/132) | 31.3% (46/147) | 34.0% (48/141) |
| 32-63 | 47.0% (62/132) | 53.1% (78/147) | 41.1% (58/141) |
| 64-127 | 9.9% (13/132) | 15.7% (23/147) | 23.4% (33/141) |
| 128-255 | 0.0% (0/132) | 0.0% (0/147) | 1.4% (2/141) |
| ≥256 | 0.0% (0/132) | 0.0% (0/147) | 0.0% (0/141) |
| day 28 after booster dose |  |  |  |
| 16-31 | 0.0% (0/122) | 0.0% (0/120) | 0.0% (0/112) |
| 32-63 | 1.6% (2/122) | 0.8 (1/120) | 0.0% (0/112) |
| 64-127 | 14.8% (18/122) | 8.3% (10/120) | 8.9% (10/112) |
| 128-255 | 35.2% (43/122) | 34.2% (41/120) | 34.8% (39/112) |
| ≥256 | 48.4% (59/122) | 56.7% (68/120) | 56.3% (63/112) |

Results expressed as n/N (%).

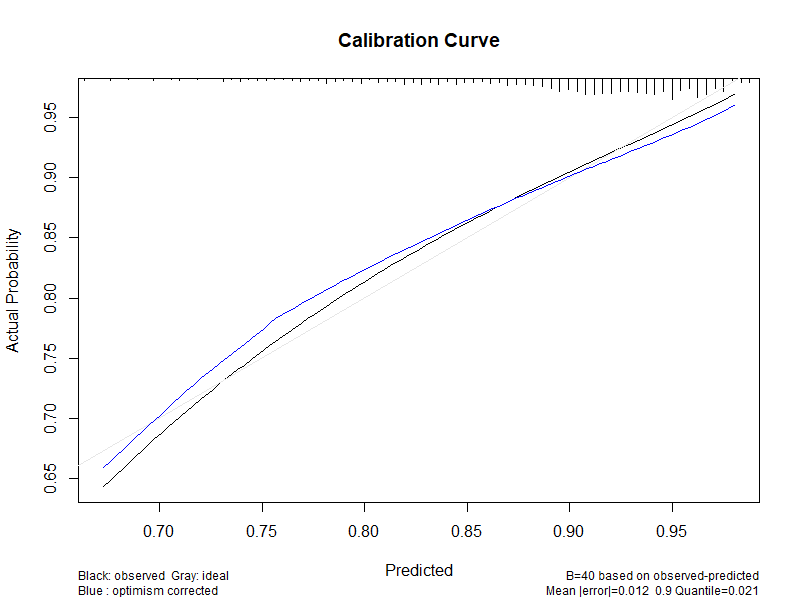

Figure S2. The calibration curves for the nomogram.

The X-axis represented the nomogram-predicted probability and Y-axis represents the actual probability of neutralizing antibody remaining positive within 10 months. Perfect prediction would correspond to the 45°gray dashed line. The black line represents the actual outcome, and the blue solid line is bias-corrected, indicating observed nomogram performance.

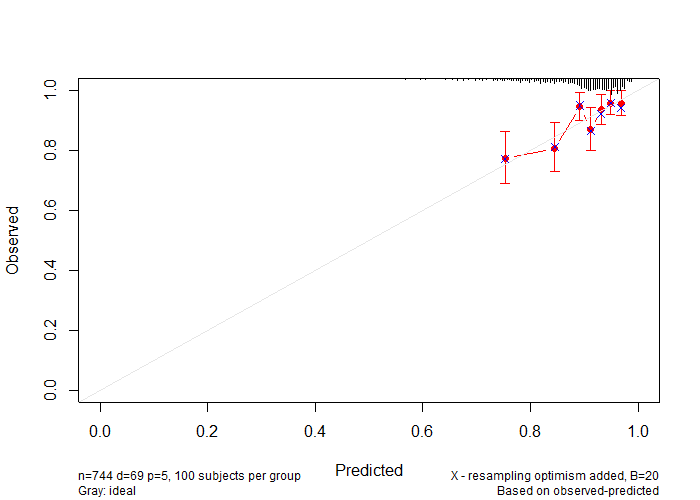

Figure S3. Calibration curve for internal verification of the prediction tool.

The calibration curves depict the calibration of the nomogram in terms of the agreement between the predicted probability of neutralizing antibody remaining positive within 10 months and observed outcomes. The 45-degree gray line represents a perfect prediction, and the pink solid lines represent the predictive performance of the nomogram. The distance between the pink solid and the ideal line represents the predictive accuracy of the nomogram.

Table S9. Summary of solicited and unsolicited adverse reactions occurred within 28 days after the third dose

| **Adverse reaction** | **0-14d-10m group**  **(n=130)** | **0-21d-10m group**  **(n=130)** | **0-28d-10m group**  **(n=130)** | ***P*** |
| --- | --- | --- | --- | --- |
| **The overall incidence of adverse reactions** | 2(1.5%) | 6(4.6%) | 4(3.1%) | 0.41^a^ |
| **Grade 1** | 2(1.5%) | 6(4.6%) | 4(3.1%) | 0.41^a^ |
| **Solicited adverse reactions within 0–7 days** |  |  |  |  |
| Any | 2(1.5%) | 4(3.1%) | 3(2.3%) | 0.91^a^ |
| Grade 1 | 2(1.5%) | 4(3.1%) | 3(2.3%) | 0.91^a^ |
| **Local reactions** | 2(1.5%) | 3(2.3%) | 2(2.3%) | 1.00 |
| Pain | 2(1.5%) | 3(2.3%) | 1(0.8%) | 0.87 |
| Grade 1 | 2(1.5%) | 3(2.3%) | 1(0.8%) | 0.87 |
| Swelling | 0(0%) | 0(0%) | 1(0.8%) | 1.00 |
| Grade 1 | 0(0%) | 0(0%) | 1(0.8%) | 1.00 |
| Pruritus | 0(0%) | 1(0.8%) | 0(0%) | 1.00 |
| Grade 1 | 0(0%) | 1(0.8%) | 0(0%) | 1.00 |
| **Systemic reactions** | 0(0%) | 1(0.8%) | 1(0.8%) | 1.00 |
| Diarrhea | 0(0%) | 0(0%) | 0(0%) | NA |
| Grade 1 | 0(0%) | 0(0%) | 0(0%) | NA |
| Fatigue | 0(0%) | 1(0.8%) | 1(0.8%) | 1.00 |
| Grade 1 | 0(0%) | 1(0.8%) | 1(0.8%) | 1.00 |
| **Unsolicited adverse reactions within 8–28 days** |  |  |  |  |
| Any | 0(0%) | 2(1.5%) | 1(0.8%) | 0.78^a^ |
| Grade 1 | 0(0%) | 2(1.5%) | 1(0.8%) | 0.78^a^ |
| **Local reactions** | 0(0%) | 1(0.8%) | 0(0%) | 1.00 |
| Rash | 0(0%) | 1(0.8%) | 0(0%) | 1.00 |
| Grade 1 | 0(0%) | 1(0.8%) | 0(0%) | 1.00 |
| **Systemic reactions** | 0(0%) | 1(0.8%) | 1(0.8%) | 1.00 |
| Cough | 0(0%) | 1(0.8%) | 0(0%) | 1.00 |
| Grade 1 | 0(0%) | 1(0.8%) | 0(0%) | 1.00 |
| Headache | 0(0%) | 0(0%) | 1(0.8%) | 1.00 |
| Grade 1 | 0(0%) | 0(0%) | 1(0.8%) | 1.00 |

Results expressed as n (%), NA=not applicable

^a^ Fisher’s exact test.

Any refers to all the participants who received booster dose with any grade adverse reactions or events.

Adverse reactions were graded according to the Guidelines for Adverse Event Classification Standards for Clinical Trials of Preventive Vaccines by the National Medical Products Administration (version 2019). Grade 1=mild; Grade 2=moderate; Grade 3=severe.

**Basic principles**

**1. Determine the curve type of the model:**

In the study of vaccine immune persistence, the commonly used curve is exponential curve. The general form of exponential curve equation is Ŷ = *k* + *α*exp (*b*X), when the independent variable x changes according to the arithmetic series, y changes according to the arithmetic series. The position, direction and curvature of the curve are determined by the constant term *k* and the positive and negative sum of parameters *α* and *b* in the equation. In practical application, the curve equation to be fitted can be determined according to the graph drawn by the measured data.

**2. Curve linearization:**

The measured data were plotted again on a semi-logarithmic graph paper (exponential curve). If the scatter has a linear trend, you can directly take the logarithm of the variable Y to find its equation; if the scatter does not show a linear trend, you can add or subtract different *k* values to X or Y and try repeatedly until the scatter is a straight line up to the trend, the obtained *k* value is the constant term *k* of the curve equation.

**3. Find the equation:**

(1) Take the natural logarithm of the independent variable X and the dependent variable Y or (Y-*k*) respectively or independently.

(2) Find the sum of squared deviations from the mean Lxx of the independent variable X, the sum of the squared deviations of the dependent variable Y, Lyy, and the product of the two variables from the mean, Lxy, Lxx and Lyy.

(3) After the above indicators are obtained, the parameter *b* can be calculated by entering the formula *b*=Lxy/Lxx, and then the value of the parameter a can be calculated by entering b into the formula LnY=*α’*+*b*X.

(4) Substituting *k*, *α’*, and *b* into the natural logarithmic form of the curve equation to be sought:

Ln (Ŷ- *k*) = ln*α*+ *b*X

(5) Restore the above formula to the exponential curve equation Y=*k* +*α* exp(*b*X).

(6) Goodness of fit analysis: calculate the determination coefficient *R^2^*, and the value range of *R^2^* is between 0 and 1; The closer *R^2^* is to 1, the better the curve fits.

**Fitting process:**

**0-14d-10m group**

1. Curve linearization: after drawing the scatter diagram on the ordinary coordinate paper according to the measured data, the curve basically conforms to the type of exponential curve; Plot the measured data on the semi logarithmic coordinate paper (exponential curve) again; The scatter point has a linear trend, so the equation can be obtained directly after taking the logarithm of the corresponding variable Y.

2. The natural logarithm of the corresponding variable Y is taken separately (Table 1).

3. Find the sum of the squares of the deviation of the independent variable X, the sum of the squares of the deviation of the dependent variable Y, and the product of the deviation of the two variables and Lxy (Table 1).

**Table 1. Calculation process of average antibody titer and curve fitting 10 months after two doses of BBIBP-CorV vaccine in 0-14d-10m group**

| **Follow-up time**  **X_1_**  **(1)** | **GMT**  **Y_1_**  **(2)** | **LnY_1_**  **(3)=Ln(1)** | **X_1_(LnY_1_)**  **(4)= (1) *(3)** | **X_1_^2^**  **(5)= (1)^2^** | **(LnY_1_)^2^**  **(6) =(3)^2^** | **Predictive value**  **Ŷ**  **(7)** | **Residual variance**  **(Ŷ-Y_1_)^2^**  **(8)** |
| --- | --- | --- | --- | --- | --- | --- | --- |
| 1 | 98.4 | 4.6 | 4.6 | 1 | 21.1 | 77.87 | 421.5 |
| 3 | 46.4 | 3.8 | 11.5 | 9 | 14.7 | 56.54 | 102.8 |
| 6 | 30.5 | 3.4 | 20.5 | 36 | 11.7 | 34.99 | 19.9 |
| 10 | 20.3 | 3.0 | 30.1 | 100 | 9.1 | 18.45 | 3.4 |
| Total 20 | 195.6 | 14.9 | 66.7 | 146 | 56.6 | NA | 547.6 |

GMT: geometric mean titers， NA=not applicable

X=5	‾Y=48.91

LnY=14.86/4=3.71

Lxx=ΣX_1_^2^-(ΣX_1_)^2^/n=146-20^2^/4=46

Lxy=66.73-(20×14.86)/4=-7.57

4. After the above indexes are obtained, substituting those into the formula *b*=Lxy/Lxx calculated parameter *b* = -7.57/46=-0.16; Then substituting *b* into the formula LnY =*α*+ *b*X to calculate the value of parameter *α*.

*b*=Lxy/Lxx=-7.57/46=-0.16

X=10 *α^’^*=3.01+0.16x=4.61

X=6	*α^’^*=3.42+0.16×6=4.38

X=3	*α^’^*=3.84+0.16×3=4.75

X=1	*α^’^*=4.59+0.16×1=4.75

The mean value of *α^’^* is 4.51

5. Substituting *α’* and *b* to the natural logarithm form of the exponential curve equation: Ln Ŷ = ln*α*+ *b*X, Ln*α*= 4.51, α=91.38

6. Restore the above formula to the exponential curve equation Ŷ =*α*exp (*b*X), then the exponential equation is: Ŷ=91.38·exp (-0.16x)

7. Goodness of fit analysis: Calculating the coefficient of determination, the closer *R^2^* is to 1, the better the curve fitting effect. The calculation formula of *R^2^* is as follows:

*R^2^*=1-Σ(Y_1_-Ŷ)^2^/Lyy

Lyy=ΣY_1_^2^-(ΣY_1_)^2^/n=13179.6909-195.63^2^/4=3611.916675

*R^2^*=1-547.613/3611.916675=0.85

8. The antibody attenuation after enhancement was predicted according to the neutralizing antibody titer 28 days after booster dose. The actual observed results (246.2) of 0-14d-10m group on the 28th day after booster dose were substituted into the index model, time 28d (calculated according to 1 month) = 1, and the calculated slope *α* values were 288.92. So, one exponential model for 0-14d-10m group was Ŷ=288.92 exp(-0.16X).

**0-21d-10m group**

1. Curve linearization: after drawing the scatter diagram on the ordinary coordinate paper according to the measured data, the curve basically conforms to the type of exponential curve; Plot the measured data on the semi logarithmic coordinate paper (exponential curve) again; The scatter point has a linear trend, so the equation can be obtained directly after taking the logarithm of the corresponding variable Y.

2. The natural logarithm of the corresponding variable Y is taken separately (Table 2).

3. Find the sum of the squares of the deviation of the independent variable X, the sum of the squares of the deviation of the dependent variable Y, and the product of the deviation of the two variables and Lxy (Table 2).

**Table 2. Calculation process of average antibody titer and curve fitting 10 months after two doses of BBIBP-CorV vaccine in 0-21d-10m group**

| **Follow-up time**  **X_1_**  **(1)** | **GMT**  **Y_1_**  **(2)** | **LnY_1_**  **(3)=Ln (1)** | **X_1_(LnY_1_)**  **(4)= (1) *(3)** | **X_1_^2^**  **(5)= (1)^2^** | **(LnY_1_)^2^**  **(6) =(3)^2^** | **Predictive value**  **Ŷ**  **(7)** | **Residual variance**  **(Ŷ-Y_1_)^2^**  **(8)** |
| --- | --- | --- | --- | --- | --- | --- | --- |
| 1 | 134.4 | 4.9 | 4.9 | 1 | 24.01 | 108.31 | 680.69 |
| 3 | 64.2 | 4.2 | 12.5 | 9 | 17.31 | 78.65 | 208.80 |
| 6 | 42.8 | 3.8 | 22.6 | 36 | 14.14 | 48.67 | 34.46 |
| 10 | 28.8 | 3.4 | 33.6 | 100 | 11.29 | 25.66 | 9.86 |
| Total 20 | 270.2 | 16.2 | 73.5 | 146 | 66.74 | NA | 933.09 |

GMT: geometric mean titers, NA=not applicable.

X=5	‾Y=67.55

LnY=16.18/4=4.05

Lxx=ΣX_1_^2^-(ΣX_1_)^2^/n=146-20^2^/4=46

Lxy=ΣX_1_(LnY_1_)-[ΣX_1_Σ(LnY_1_)]/n=73.54-(20×16.18)/4=-7.36

4. After the above indexes are obtained, substituting those into the formula *b*=Lxy/Lxx calculated parameter -7.36/46=-0.16; Then substituting *b* into the formula LnY =*α*+ *b*X to calculate the value of parameter α.

*b*=Lxy/Lxx=-7.36/46=-0.16	LnY=*α^’^* +*b*x

X=10	*α^’^*=3.36+0.16×10=4.96

X=6	*α^’^*=3.76+0.16×6=4.72

X=3	*α^’^*=4.16+0.16×3=4.64

X=1	*α^’^*=4.9+0.16×1=5.06

The mean value of *α^’^* is 4.85

5. Substituting *α’* and *b* to the natural logarithm form of the exponential curve equation: Ln Ŷ = ln*α*+ *b*X, Ln*α*=4.85, *α=*127.10

6. Restore the above formula to the exponential curve equation y =*α*exp (*b*X), then the exponential equation is: Ŷ=127.10 exp (-0.16x)

7. Goodness of fit analysis: Calculating the coefficient of determination, the closer *R^2^* is to 1, the better the curve fitting effect. The calculation formula of *R^2^* is as follows:

*R^2^*=1-Σ(Y_1_-Ŷ)^2^/Lyy

Lyy=ΣY_1_^2^-(ΣY_1_)^2^/n=24846.28-270.2^2^/4=6594.27

*R^2^*=1-933.8071/6594.27=0.86

8. The antibody attenuation after enhancement was predicted according to the neutralizing antibody titer 28 days after booster dose. The actual observed results (277.5) of the three groups on the 28th day after booster dose were substituted into the index model, time 28d (calculated according to 1 month) = 1, and the calculated slope *α* values were 325.65. So, one exponential model for the 0-21d-10m group was Ŷ=325.65 exp(-0.16x).

**0-28d-10m group**

1. Curve linearization: after drawing the scatter diagram on the ordinary coordinate paper according to the measured data, the curve basically conforms to the type of exponential curve; Plot the measured data on the semi logarithmic coordinate paper (exponential curve) again; The scatter point has a linear trend, so the equation can be obtained directly after taking the logarithm of the corresponding variable Y.

2. The natural logarithm of the corresponding variable Y is taken separately (Table 3).

3. Find the sum of the squares of the deviation of the independent variable X, the sum of the squares of the deviation of the dependent variable Y, and the product of the deviation of the two variables and Lxy (Table 3).

**Table 3. Calculation process of average antibody titer and curve fitting 10 months after two doses of BBIBP-CorV vaccine in 0-28d-10m group**

| Follow-up time  X_1_  (1) | GMT  Y_1_  (2) | LnY_1_  (3)=Ln (1) | X_1_(LnY_1_)  (4)= (1) *(3) | X_1_^2^  (5)= (1)^2^ | (LnY_1_)^2^  (6) =(3)^2^ | Predictive value  Ŷ  (7) | Residual variance  (Ŷ-Y_1_)^2^  (8) |
| --- | --- | --- | --- | --- | --- | --- | --- |
| 1 | 145.5 | 5.0 | 5.0 | 1 | 24.8 | 118.27 | 741.5 |
| 3 | 71.5 | 4.3 | 12.8 | 9 | 18.2 | 86.40 | 222.0 |
| 6 | 47.1 | 3.9 | 23.1 | 36 | 14.8 | 53.95 | 46.9 |
| 10 | 32.4 | 3.5 | 34.8 | 100 | 12.1 | 28.79 | 13.0 |
| Total 20 | 296.5 | 16.6 | 75.7 | 146 | 70.0 | NA | 1023.4 |

GMT: geometric mean titers, NA=not applicable.

‾X=5	‾Y=74.13

LnY=16.58/4=4.15

Lxx=ΣX_1_^2^-(ΣX_1_)^2^/n=146-20^2^/4=46

Lxy=ΣX_1_(LnY_1_)-[ΣX_1_Σ(LnY_1_)]/n=75.69-(20×16.58)/4=-7.21

4. After the above indexes are obtained, substituting those into the formula *b*=Lxy/Lxx calculated parameter *b* = -7.21/46=-0.157; Then substituting b into the formula LnY =*α*+ *b*X to calculate the value of parameter *α*.

*b*=Lxy/Lxx=-7.21/46=-0.157	LnY=*α^’^* +*b*x

X=10	*α^’^*=5.05

X=6	*α^’^*=4.79

X=3	*α^’^*=4.74

X=1	*α^’^*=5.14

The mean value of *α^’^* is 4.93

5. Substituting *α’* and *b* to the natural logarithm form of the exponential curve equation: Ln Ŷ = ln*α*+ *b*X, Ln*α*= 4.93, *α*= 138.38

6. Restore the above formula to the exponential curve equation y =*α*exp (*b*X), then the exponential equation is: Ŷ=138.38·exp (-0.16x)

7. Goodness of fit analysis: Calculating the coefficient of determination, the closer *R^2^* is to 1, the better the curve fitting effect. The calculation formula of *R^2^* is as follows:

*R^2^*=1-Σ(Y_1_-Ŷ)^2^/Lyy

Lyy=ΣY_1_^2^-(ΣY_1_)^2^/n=29550.67-296.5^2^/4=7572.61

*R^2^*=1-1023.43/7572.6075=0.86

8. The antibody attenuation after enhancement was predicted according to the neutralizing antibody titer 28 days after booster dose. The actual observed results (288.6) of the three groups on the 28th day after booster dose were substituted into the index model, time 28d (calculated according to 1 month) = 1, and the calculated slope *α* values were 338.67 respectively. So, one exponential model for the 0-14d-10m group was Ŷ=338.67·exp (-0.16x).
