## Supplementary material for "Long-term immune persistence induced by two-dose BBIBP-CorV vaccine with different intervals, and immunogenicity and safety of a homologous booster dose in high-risk occupational population Secondary Study Based on a Randomized Clinical Trial": protocol 1

### 随机、对照、多中心疫苗试验对新型冠状病毒灭活疫苗不同免疫程序的安全性和免疫原性观察研究项目委托协议书

甲方：　北京生物制品研究所有限责任公司

地址：　北京市北京经济技术开发区博兴二路9号院

联系人：王明

联系电话：010-60963524

乙方：　山西省疾病预防控制中心

地址：　山西省太原市小南关街8号

联系人：张晓红

联系电话：0351-7553368

北京生物制品研究所有限责任公司（甲方）研制的"新型冠状病毒灭活疫苗（Vero 细胞）"，依据国家有关规定，特委托山西省疾病预防控制中心（乙方）作为该疫苗临床研究负责机构，开展随机、对照、多中心疫苗试验对新型冠状病毒灭活疫苗不同免疫程序的安全性和免疫原性观察研究项目工作。甲乙双方经协商达成如下协议：

**一、研究的基本情况：**

（一）研究项目：随机、对照、多中心疫苗试验对新型冠状病毒灭活疫苗不同免疫程序的安全性和免疫原性观察研究

（二）产出成果：随机、对照、多中心疫苗试验对新型冠状病毒灭活疫苗不同免疫程序的安全性和免疫原性评价报告

**二、双方的责任和义务：**

（一）甲方的责任和义务：

1.负责制定研究方案和提供临床研究用经费；

2.对项目研究期间受试者发生的不良事件，协助乙方对受试者进行救治，并积极配合对不良事件的调查、诊断和处置工作，负责落实疫苗生产企业按有关规定承担相关的补偿费用；

3.协助乙方完成项目所需的伦理审批，提供相应资料；

4.完成其他经双方协商应由甲方承担的工作。

（二）乙方的责任和义务：

1.负责选定研究现场并组织和实施工作；

2.严格按照研究方案要求进行研究，确保临床研究的质量；

3.负责与伦理审查委员会沟通，向伦理审查委员会提供伦理审查需要

的所有文件，同时负责获得和保存伦理审查委员会下发的伦理审查意见以及伦理审查相关的其他记录和文件；

4. 负责督导、协调现场完成受试者知情同意、接种、随访和观察等工作；

5. 负责督导现场生物标本采集、处理、送检及安全性观察等工作；

6. 负责对受试者研究期间出现的不良事件进行调查诊断并记录在案；

7. 保证将研究数据真实、准确、完整、及时地录入病例报告表；

8. 负责在规定的时间内完成现场小结的撰写；

9. 负责按项目进度定期向甲方提供项目进展情况汇报；

10. 负责撰写符合要求的研究总结报告；

11. 完成其他经双方协商应由乙方承担的工作。

**三、实施时间**

本临床研究自 2020 年 10 月开始，整个临床研究项目持续约 18 个月，期间完成 732 名受种者入组、样本采集、随访、免疫原性及安全性评价现场工作。完成时间如有变动，双方另行协商。

**四、研究经费、支付方式和支付单位：**

（一）观察研究经费：甲方向乙方提供临床研究经费合计人民币 170 万元（大写：壹佰柒拾万元整）（含税），用于支付与本项目研究有关的各项工作费用。

（二）支付方式：分两次支付，第一次在协议签订后 15 个工作日内支付 119 万元，第 2 次在乙方按照方案要求完成全部现场工作并提交研究总结报告后支付人民币 51 万元。

（三）乙方在收到项目经费后负责提供合规的发票，内容“临床观察费”。

（四）乙方管理和使用研究经费应符合国家相关政策及规定。

**五、不可抗力**

（一）协议中任何一方，由于严重自然灾害和相关疾病的流行等不可预见且无法抗拒的客观情况而影响协议执行时，可延长履行协议的期限，延长时间相当于事故所影响的时间。发生事故一方应在事件发生48 小时内将发生不可抗力事件的情况通知协议他方，并及时将有关证明文件提交他方确认。因政府原因造成本协议终止的，比照本款约定执行。

（二）如果在本协议执行过程中因政府、监管部门的原因造成协议部分或全部工作不能按时完成的，双方同意工作完成期限顺延，且双方均不就因此产生的逾期承担违约责任。

（三）如果本协议或临床研究被提前终止，乙方应尽最大努力减少因提前终止协议或临床研究而造成的损失。非因协议任一方原因导致本协议终止的，双方协商确定费用结算事宜。

**六、违约责任：**

（一）本协议正式签订后，任何一方不履行或不完全履行本协议约定条款的，即构成违约。协议一方违反本合同规定，造成对方经济损失的，守约方有权要求解除本合同，并由违约方承担赔偿责任。

（二）甲方必须按本协议约定的付款进度及时支付相应款项。若不能按时支付，则按照每延迟一天支付应付款项金额的 0.25%作为违约金给乙方（因乙方原因而延迟付款的时间不在此列）。违约金最高限额是应付款项的 5%，应付违约金累计达到或超过最高限额，乙方有权解除合同，除上述违约金外，还有权要求甲方按 5%另行支付违约赔偿。

（三）因乙方原因造成的本协议提前解除的，乙方应向甲方退还未发生的费用，对于已发生的费用，按照实际价格计算。因甲方原因造成本协议提前解除的，乙方对甲方已付费用不予退还，此外对本试验阶段已发生的费用，甲方应按照乙方实际支出进行赔付。

**七、承诺和保证**

（一）除获得书面同意外，任何一方不得将对方的名称或其他任何信息用于任何与本协议无关的任何事项或活动中，也不得声称或以任何方式直接或间接使他方误解任何双方存在除本协议约定以外的任何关系。否则，应承担因此产生的全部责任。

（二）任何一方对于因履行本协议而获知或取得的对方的单位信息、资料、图表或以电子文件形式存在的保密信息负有保密的义务，未经保密信息所有者的书面同意，不得擅自泄露、使用或许可第三方使用，法律、法规另有规定的除外。

**八、成果归属及保密：**

（一）本协议项下研究项目成果归甲乙双方共同所有。项目成果可由双方在研讨会、全国性或区域性专业会议上，或是在刊物、论文中，或是以其他方式，对该项目研究的方法和成果进行介绍、发表和公布。

（二）双方均承诺对研究项目涉及的专有技术及相关资料严格保密。

**九、争议的解决方式：**

发生与本协议书有关的一切争议，应首先友好协商解决。如果协商解决不成，双方均同意提交乙方所在地人民法院诉讼解决。

**十、附则：**

（一）本协议书自双方签字、盖章之日起生效。

（二）本协议书一式四份，甲乙双方各执两份，同等有效。

（三）本协议书签订后，如需修改，经双方协商一致签订补充协议或工作备忘录并作为本协议之组成部分，与本协议共同使用。附件为本协议的组成部分，与本协议具同等法律效力。

（四）双方确认的联系方式及账户信息：

1、甲方通讯地址：北京市北京经济技术开发区博兴二路 9 号院 4 号楼 2 层 205 室邮编：100000 电话：010-60963524 、60963038

传真：010-60963311 联系人：王明

名称：北京生物制品研究所有限责任公司

纳税人识别号：91110 302MA 007H9 5X1

账号：0200006809200111871

开户行：中国工商银行股份有限公司北京管庄支行

2、乙方通讯地址：山西省太原市小南关街 8 号邮编：030000 电话：0351-7553368 联系人：张晓红

账户名称：山西省疾病预防控制中心

开户行：140451529895

账　号：中国银行太原北城支行

任何与履行本协议书有关的文件均应以书面形式送达，送达地址以本协议第十条第（四）款确认的地址为准。若任一方变更联系地址（即送达地址）应向对方提交书面变更函，若未提交，任一方向本协议约定的地址邮寄通知等材料，均视为送达。

甲方：北京生物制品研究所有限责任公司

法定代表人：王辉

或委托代理人签字：

签署日期：2021年9月23日

乙方：山西省疾病预防控制中心

法定代表人：

或委托代理人签字：陈靖

签署日期：2021年9月23日
