## Supplementary material for "Long-term immune persistence induced by two-dose BBIBP-CorV vaccine with different intervals, and immunogenicity and safety of a homologous booster dose in high-risk occupational population Secondary Study Based on a Randomized Clinical Trial": protocol 2

**Protocol Title: A randomized, controlled, multicenter study on the safety and immunogenicity of SARS-CoV-2 inactivated vaccine**

**Initial study**

Objectives of Study：

To evaluate the immunogenicity and safety of the SARS-CoV-2 inactivated vaccine at the different schedule of day 0, 14, day 0, 21, or day 0, 28.

Participants

We conducted a randomized, controlled phase IV trial of the SARS-CoV-2 inactivated vaccine manufactured by Beijing Biological Products Institute Co., Ltd. between January and May 2021 in Taiyuan City, Shanxi Province, China. Eligible participants were public security officers and airport ground staff aged 18–59 years. Written informed consent was obtained from all participants before the enrolment in the initial study. The clinical trial protocol for the study was approved by the Ethics Committee of Shanxi Provincial Center for Disease Control. The study was done in accordance with the Declaration of Helsinki and Good Clinical Practice. The trial was registered with ChiCTR.org.cn (ChiCTR2100041705, ChiCTR2100041706).

Inclusion Criteria:

1. Subjects aged 18-59 years old;

2. Without previous SARS-CoV-2 vaccination and infection;

3. Negative for SARS-CoV-2 nucleic acid.

Exclusion Criteria:

1. History or family history of allergy, convulsion, epilepsy, encephalopathy or psychosis;

2. Any intolerance or allergy to any component of the vaccine;

3. known or suspected diseases including severe respiratory disease, severe cardiovascular disease, severe liver or kidney disease, medically uncontrollable hypertension (systolic blood pressure ≥ 140mmHg and diastolic blood pressure ≥ 90mmHg), complications of diabetes mellitus, malignancy, various acute diseases or acute episodes of chronic disease;

4. Various infectious, suppurative and allergic skin diseases; congenital or acquired immunodeficiency;

5. Other vaccination history within 14 days before vaccination;

6. A history of coagulation dysfunction, a history of non-specific immunoglobulin injection within 1 month prior to enrollment; acute illness with fever (body temperature > 37.0°C);

7. Being pregnant or breastfeeding.

Sample size

The study sample size of 360 participants provided 84.4% power to detect a difference of 5% (85% vs 80%) of responders in the 0-21 and 0-28 vaccination groups compared with the 0-14 group, respectively in public security officers and airport ground staff.

Baseline questionnaire

Demographic information [age, sex, body mass index (BMI), marital status, and education level], influenza vaccination history, smoking, drinking, and chronic diseases were collected via questionnaire investigation.

Randomized

A computerized random number generator performed block randomization with a randomly selected block size of 6, and eligible participants were randomly assigned into three groups to receive two doses inactivated SARS-CoV-2 vaccine at the schedule of day 0-14, day 0-21, or day 0-28.

Vaccination

Each dose of vaccine containing 4 µg of inactivated SARS-CoV-2 virus antigen was intramuscularly injected into the lateral deltoid muscle of the upper arm. The vaccines used in this study were inactivated vaccine (Vero Cell) produced by Beijing Biological Products Institute Co., Ltd.

Follow-up of priming two-dose vaccination

A total of 256, 247 and 241 patients in the 0-14, 0-21, and 0-28 groups, respectively, completed the follow-up at day 28 after the second dose vaccination.

The contents of follow-up included the blood samples, oropharyngeal/nasal swabs, and adverse reactions collection.

Laboratory methods

Oropharyngeal/nasal swabs were collected for detecting SARS-CoV-2 nucleic acid from all subjects by using reverse transcriptase-polymerase chain reaction (RT-PCR) test. Blood samples were taken from participants for serology tests before the first injection and on day 28 after the second injection. The neutralizing antibody to live SARS-CoV-2 [strain 19nCoV-CDC-Tan-Strain 05 (QD01)] were quantified using a micro cytopathogenic effect assay at baseline and 28 days after immunization. A positive antibody response (seroconversion) was defined as post-injection titer of at least 1:16 if the baseline titer was below 1:4, or at least a fourfold increase in post-injection titer from baseline if the base-line titer was at least 1:4. We defined the neutralizing antibody seroconversion rate as post-injection titer of a 16-fold (Baseline titers were all below 1:4).

Statistical analysis

Data were recorded using EpiData version 3.1 (EpiData Association, Odense, Denmark), and analyses were performed using SAS version 9.3 (SAS Institute, Cary, NC, USA). Analysis of variance (ANOVA) was used to analyze continuous data, and the chi-square or Fisher’s exact test was used for categorical data. We assessed immunogenic end-points by the intention-to-treat (ITT) analysis (i.e., subjects who undertake randomization) and per-protocol (PP) analysis (i.e., subjects who compliant to the protocol, receive 2 doses of vaccine according to the requirements of the protocol, and have serum-testing results before and after immunization). Multinomial logistic regression analysis and unconditional logistic regression model were used to determine the influencing factors of SARS-CoV-2 neutralizing antibody immunization. The safety analysis was performed on data from all subjects who received vaccination after randomization. The level of statistical significance for all analyses was *P* < 0.05.

Safety assessment

After each dose was vaccinated, the participants were observed for any immediate reaction for 30 min, and local and systemic adverse reactions were collected. Participants were required to record the local adverse events and systemic adverse events on diary cards within 7 days of each injection. Any other unsolicited symptoms were also recorded during a 28-day follow-up period after each injection by spontaneous report from the participants combined with the regular visit. The solicited adverse reactions included local reactions (pain, induration, swelling, rash, flush, and pruritus) and systematic reactions [fever, diarrhea, dysphagia, anorexia, vomiting, nausea, muscle pain (non-vaccination sites), arthralgia, headache, cough, dyspnea, skin and mucosal abnormalities, acute allergic reactions, and fatigue].

Outcomes

The primary immunogenic endpoints were the seroconversion rates [geometric mean titer (GMT) ≥ 16] and GMT of neutralizing antibody to live SARS-CoV-2 at day 28 after the last dose.

Secondary immunogenic endpoints were the positive rates (GMT ≥ 32, 64, 128, 256) 28 days after the whole course of vaccination, respectively.

The primary endpoint for safety was the occurrence of adverse reactions within 7 days after the first and second vaccinations.

Secondary safety endpoints were adverse events within 28 days after the first and the second vaccinations across the three groups.

Secondary study

Part I: Long-term immune persistence

Objectives of Study：

To assess the immune persistence of three two-dose schedules of either 14 days, 21 days or 28 days of BBIBP-CorV vaccine based on the initial study.

Participants

The responders at day 28 after priming two-dose vaccine were followed up at months 3, 6 and 10. Written informed consent was obtained from all participants before the enrolment in the initial study. The clinical trial protocol for the study was approved by the Ethics Committee of Shanxi Provincial Center for Disease Control. The study was done in accordance with the Declaration of Helsinki and Good Clinical Practice. The trial was registered with ChiCTR.org.cn (ChiCTR2100041705, ChiCTR2100041706).

Follow-up of priming two-dose vaccination

We followed up participants who were antibody positive (GMT≥16) at day 28 after the second dose. Of them were followed up at months 3, 6 and 10 to evaluate the immune persistence of three two-dose schedules.

The contents of follow-up included the blood samples, oropharyngeal/nasal swabs, and adverse reactions collection.

Laboratory methods

Oropharyngeal/nasal swabs for RT-PCR (reverse transcriptase-polymerase chain reaction) testing were collected from all participants at each follow-up point.

The neutralizing antibody to live SARS-CoV-2 (strain 19nCoV-CDC-Tan-Strain 05 [QD01]) were quantified using a micro cytopathogenic effect assay at 3, 6 and 10 months after the second dose, and 28 days after the booster dose. The lower limit of detection was 4 for the neutralizing antibody test. We defined positive antibody response as a titer of 16 or greater for neutralizing antibodies to infectious SARS-CoV-2. We assessed the positive rates of neutralizing antibody different criteria [geometric mean titers (GMT) ≥ 16, 32, 64, 128 or 256].

Part II: booster dose vaccination

Objectives of Study：

To evaluate the immunogenicity and safety of booster dose at month 10 after the second dose.

Participants

A total of 390 participants who met the inclusion and exclusion criteria were received the booster dose 10 months after the second dose.

Written informed consent was obtained from all participants before the enrolment in the initial study. The clinical trial protocol for the study was approved by the Ethics Committee of Shanxi Provincial Center for Disease Control. The study was done in accordance with the Declaration of Helsinki and Good Clinical Practice. The trial was registered with ChiCTR.org.cn (ChiCTR2100041705, ChiCTR2100041706).

Inclusion Criteria:

1. No an unacceptable adverse event;

2. No SARS-CoV-2 infection during the follow-up period;

3. Completed the initial two doses of vaccination;

4. Be able and willing to receive the booster dose injection.

Exclusion Criteria:

1. A COVID-19 vaccine other than the experimental vaccine was used during the study period;

2. Acute or new chronic disease occurs during the study period, which including severe respiratory disease, severe cardiovascular disease, severe liver or kidney disease, medically uncontrollable hypertension (systolic blood pressure ≥ 140 mmHg and diastolic blood pressure ≥ 90mmHg), complications of diabetes mellitus, malignancy, various acute diseases or acute episodes of chronic disease;

3. Other vaccination history within 14 days before vaccination;

4. Being pregnant or breastfeeding before vaccination;

5. According to the judgment of the investigator, participants had any factors not suitable for vaccination.

Booster vaccination

At month 10, those participants who met the inclusion and exclusion criteria were received a homologous booster dose based on three two-dose schedules, abbreviated as 0-14d-10m, 0-21d-10m and 0-28d-10m group.

The vaccines were administered intramuscularly in the deltoid region of the upper arm with a dosage of 4µg. The vaccines used in this study were Beijing Institute of Biological Products Co., Ltd. (‎BIBP)‎ COVID-19 Vaccine, by China National Biotec Group (‎CNBG)‎, Sinopharm (‎international non-proprietary name: Covilo; BIBP-CorV)‎

Follow-up after booster dose vaccination

The participants were followed-up at day 28 after booster dose for assessing the booster-induced immunogenicity and safety. The contents of follow-up included the blood samples, oropharyngeal/nasal swabs, and adverse reactions collection.

Laboratory methods

Oropharyngeal/nasal swabs for RT-PCR (reverse transcriptase-polymerase chain reaction) testing were collected from all participants at each follow-up point.

The neutralizing antibody to live SARS-CoV-2 (strain 19nCoV-CDC-Tan-Strain 05 [QD01]) were quantified using a micro cytopathogenic effect assay at 3, 6 and 10 months after the second dose, and 28 days after the booster dose. The lower limit of detection was 4 for the neutralizing antibody test. We defined positive antibody response as a titer of 16 or greater for neutralizing antibodies to infectious SARS-CoV-2. We assessed the positive rates of neutralizing antibody different criteria [geometric mean titers (GMT) ≥ 16, 32, 64, 128 or 256].

Statistical analysis

We calculated the GMT, positive rates and the constituent ratio of neutralizing antibodies at day 28 post-booster. Safety endpoints were presented descriptively as frequencies (%) per group. Analysis of Variance (ANOVA) was used to analyze neutralizing antibody levels of different groups which were log-transformed. Chi-square test or Fisher precision test was used to Categorical data.

An exponent curve model was used to predict the neutralizing antibody levels after the booster dose, and the general form of exponential curve equation is Ŷ = *k* + *α* exp (*b*X). The position, direction and curvature of a curve are determined by the sign and magnitude of constant terms and parameters *α* and *b*, respectively. The actual antibody levels (Y) over time (X) of follow-up from 28 days (calculated according to 1 month) to 10 months after two doses were used to construct the model. After the actual values were transformed into logarithm, reciprocal or square root, the curve equation was linearized, and the least square method was used to fit the optimal exponential model of vaccine immune persistence. Based on the model, the antibody attenuation after enhancement was predicted according to the actually observed neutralizing antibody titer decay after two doses. The values of *P*<0.05 were considered statistically significant. Statistical computations were performed using SAS version 9.4 and graphing performed using GraphPad Prism 9.0.0.

Safety assessment

Safety information after the booster dose was obtained by the same methods as for the first two doses. Participants were observed for 30 minutes in the observation room after booster dose vaccination for any acute reactions. Participants were instructed to record local and systemic reactions daily for 7 days on diary cards. Solicited local adverse events included pain at the injection site, induration and swelling, and systemic adverse events included diarrhea and dysphagia. For days 8-28, participants spontaneously reported any unsolicited adverse events including local reactions (rash) and Systemic reactions (cough and headache). Adverse reactions were graded according to the Guidelines for Adverse Event Classification Standards for Clinical Trials of Preventive Vaccines by the National Medical Products Administration (version 2019). Grade 1=mild; Grade 2=moderate; Grade 3=severe.

Outcomes

Secondary immunogenic endpoints: GMT of neutralizing antibodies to live SARS-CoV-2, the positive rates of different criteria and the constituent ratio of GMT of neutralizing antibodies(GMT 16-31，GMT 32-63， GMT 64-127， GMT 128-255 and GMT ≥ 256) at 28 days after booster dose.

Exploratory immunogenic endpoints: the prediction of neutralizing antibody decay after the booster dose.

The primary safety endpoints: adverse events within 7 days after booster dose. Secondary safety endpoints: any adverse events within 28 days after the booster dose vaccinations across the three groups.
